# Pediatric Norovirus Vaccination: Modeling the Influence of Schedule on Population Health

**DOI:** 10.1101/2024.04.26.24305745

**Authors:** Elizabeth Sajewski, Alicia NM Kraay, Juan S Leon, Katia Koelle, Andreas Handel, Ben A Lopman

## Abstract

**Introduction:** Noroviruses cause approximately 20 million cases of acute gastroenteritis (AGE) annually in the United States, with children under 5 years of age experiencing the highest incidence. With promising norovirus vaccines moving into pivotal trials in children, an important next step in norovirus vaccine evaluation is comparing benefits of various pediatric vaccination schedules.

**Methods:** We developed an age-structured deterministic compartmental model for norovirus transmission and fit it to norovirus case incidence data representative of the United States population. The model accounted for immunity accrued across multiple infections and potential maternal antibody interference with vaccine performance. Using this model, we simulated five pediatric vaccination schedules (at 0/2/4m, 2/4/6m, 6/12m, 12/24m, and 48m) with 70% and 90% vaccination coverage, with and without maternal antibody interference. We evaluated the impacts of vaccination on clinical outcomes, including acute gastroenteritis cases, outpatient visits, hospitalization, and death due to norovirus.

**Results:** Vaccination impacts were primarily concentrated in the pediatric age group, with modest indirect effects to unvaccinated age groups. Comparing scenarios, the 2/4/6m schedule was more impactful, with a 43% (95% uncertainty interval (UI): 29 – 62%) reduction in pediatric AGE cases (<5 years old) and 19% (95% UI: 7 – 54%) reduction in total population cases (including pediatric cases) with 70% vaccination coverage and no maternal antibody interference. This scenario was estimated to avert 12,200 (95% US: 7,100 – 21,500) AGE cases, 2,100 (900 – 4,500) outpatient visits, and 50 (30 – 90) hospitalizations per 100,000 young children. In the total population, 3.72 million (95% UI: 2.06 – 7.07 million) AGE cases, 526,000 (24,800 – 1,092,000) outpatient visits, 18,000 (9,500 – 41,600) hospitalizations, and 152 (52-602) deaths were estimated to be averted across the population, with 38% of the overall impact conferred by indirect effects. While the 0/2/4m schedule had the greatest overall impact when assuming no interference from maternal antibody, the 2/4/6m schedule was less affected by interference from maternal antibodies. Assuming maternal antibody interference does compromise vaccine efficacy, the 2/4/6m schedule was comparable to the most impactful vaccination schedule with maternal antibody interference (6/12m schedule, 16% [95% UI: 7 – 29%] of pediatric cases and 6% [95% UI: 1 – 26%] of total cases averted, 2/4/6m schedule: 14% [95% UI: 7 – 25%] of pediatric cases and 6% [95% UI: 2 -21%] of total cases averted). Thus, while the total vaccination impact was highly sensitive to assumptions regarding maternal antibody interference, the 2/4/6m schedule impacts were robust across assumptions.

**Significance:** Pediatric norovirus vaccination scheduled at 2/4/6m and similar schedules have the potential to substantially reduce norovirus burden in high incidence age groups and avert healthcare utilization, likely with modest benefits to unvaccinated individuals.

## Introduction

Globally, among all age groups, norovirus is the leading viral cause of acute gastroenteritis (AGE). Norovirus infection annually leads to between 140 and 667 million AGE cases and between 71,000 and 212,000 deaths^1–6^. In the United States (US), across all age groups, norovirus infection leads to 570 – 800 deaths, 56,000 – 71,000 hospitalizations, 400,000 emergency department visits, 1.7 – 1.9 million outpatient visits, and 19 – 21 million total illnesses ^7–10^. Norovirus also causes a substantial economic burden, with hospitalizations in the US alone estimated to cost approximately $500 million annually^9^. In areas where rotavirus vaccination has been implemented, including the US, norovirus is also the leading cause of medically attended AGE in pediatric populations ^11–14^. With the significant health, economic, and societal burdens of norovirus, pediatric norovirus vaccination may be an important tool in reducing morbidity and mortality in children and across the population^1,3,4,8,15^.

There are several norovirus vaccine candidates currently under development^16^. Furthest along is a bivalent GI.1/GII.4 virus-like particle (VLP) intramuscular product which has completed Phase II trials in children (6 weeks to 8 years of age), as well as adults (18 – 50 years of age) and older adults (60+ years of age). Trials have demonstrated safety and immunogenicity of multi-dose bivalent norovirus VLP candidates in children and adults, with greater seroconversion rates among children^16–19^. There is limited efficacy data in adults but none yet in children.

Both children and older adults have been considered as potential target groups for norovirus vaccination. While older adults experience the highest incidence of severe outcomes (i.e., death), children under 5 years of age experience the highest incidence of subclinical and medically-attended norovirus AGE ^6,15,20^. Additionally, children are likely to be more infectious than adults and play an outsized role in transmission to all age groups^21^. Therefore, pediatric vaccination may have the greatest direct and indirect benefits compared to older adult vaccination. This hypothesis is supported by previous modeling work which indicated that pediatric norovirus vaccination could avert ∼60% of norovirus AGE and health care utilization among children under 5 years of age as well as have considerable indirect effects reducing norovirus burden across all age groups^22^. Specifically, the indirect effects of pediatric vaccination resulted in norovirus reductions in older adults comparable to directly vaccinating the older population^22^. Pediatric, compared to older age groups, norovirus vaccination is also predicted to provide the most economic value, with a more favorable ‘cost per case averted’ estimate^23^.

When considering pediatric norovirus vaccination strategies, ideally, a child would be protected for as long as they remain at high risk of infection and severe disease. To achieve the greatest protection, the optimal age of immunization may be some time after birth for several reasons. First, initially after birth, children may be protected against norovirus by maternal immunity^24,25^. For norovirus, passively-acquired maternal antibodies wane over the first 6 months of life^24^. These maternal antibodies likely provide passive immunity against norovirus and contribute to the lower incidence of norovirus observed among children under 6 months of age compared to children 6 to 24 months of age^26^. Maternal antibodies may also interfere with norovirus vaccine immunogenicity in infants. Though the degree of maternal antibody interference for norovirus vaccine immunogenicity specifically is unclear, no pediatric vaccines among all recommended pediatric vaccinations are fully effective in the presence of maternal antibodies. The limited effect of vaccines in children with maternal antibodies is potentially due to inhibition of the generation of neutralizing antibodies^27^. This effect is well studied for measles, where vaccination is delayed until one year of age in the US to allow for measles-specific maternal antibodies to wane^27^. Similarly, rotavirus vaccine immunogenicity is inhibited by maternal antibodies, with higher levels of maternal antibodies in infants associated with reduced vaccine seroconversion^28^. Accordingly, there may be a tradeoff between the protection potentially provided by early vaccination, when the risk of severe outcomes is highest, with the potential for maternal antibody interference with vaccine immunogenicity.

Second, children experience the highest incidence of norovirus AGE between 6 and 24 months of age^26,29^, suggesting that protection from norovirus may be most impactful during this period. Given that natural immunity to norovirus is not lifelong^21,30^, vaccine-derived immunity is likely to also wane.. Accordingly, the age of immunization should be optimized to protect children for the duration of this age range of highest risk from norovirus AGE between 6 and 24 months of age.

Third, primary norovirus infections may provide limited immunity in children, with protection building over subsequent infections^31^. Analysis of birth cohorts have found that young children with two or more prior norovirus infections have increased protection against norovirus infection and, to a greater extent, disease, compared to children with one or no prior norovirus infections^31^. These analyses indicate that norovirus immunity may be accumulated over several infections in young children^31^ These early life dynamics and building of partial immunity in infants may factor into schedules for the optimal time of initial and subsequent doses in a multi-dose pediatric norovirus vaccination series.

The objective of this study is to compare the population-level impacts of five pediatric norovirus immunization schedules that align with existing pediatric vaccination schedules recommended by the US Advisory Committee on Immunization Practices (ACIP). We developed an age-structured dynamic norovirus transmission model, building on previous studies by incorporating key early life infection dynamics, including maternal immunity and immune protection accumulated over multiple norovirus infections early in life. We used a detailed dataset with fine pediatric age strata to fit the compartmental transmission model. We then simulated the impacts of different pediatric vaccination schedules under realistic vaccine coverage scenarios in the presence and absence of maternal antibody interference with vaccination immunogenicity. We calculated direct and indirect impact of norovirus vaccination in specific age groups and in the total US population across four outcomes: AGE cases, outpatient visits, hospitalizations, and deaths.

## Methods and data

### Data

With the aim of capturing norovirus transmission and simulating the impact of vaccination on the US population, we fit a dynamic compartmental transmission model to a nine-year time series of age-stratified norovirus AGE cases from Germany from 2011-2019. This dataset included a total of 1.3 million lab-confirmed cases reported to SurvStat, a notifiable disease database collected by the Robert Koch Institute^32^. Data were reported in single-year age groups, which provided detailed pediatric norovirus AGE case counts. Such detailed data were not available for the US, so we made the assumption that age-specific incidence in the US would be similar to Germany based on similar income level and demographic structure.

We further stratified infant (<1 year of age) incidence into two 6-month age groups, with the apportionment based on meta-analysis results of developed country age distributions of pediatric norovirus AGE cases^26^. We then standardized the German data set to the US population using age-specific population ratios from CDC Wonder^33^ and UN World Population Prospects^34^. We established a reporting rate based on the standardized case report data and the expected total number of norovirus AGE cases across all age groups in the US (∼20 million)^8^, such that the estimated reporting rate scaled the case report data to the expected total number of cases in the US.

### Model Design

We developed a deterministic, age-structured dynamic compartmental transmission model building on our and others’ previously published norovirus vaccination models combined with elements of a pediatric rotavirus vaccination model^22,35–37^. To simulate different pediatric immunization schedules, this model tracks transmission in finer age groups than previous models (Figure 1). This model differs from previous models used to evaluate norovirus vaccination strategies^22,38^ in that it allows immunity to build over multiple infections, consistent with observations form birth cohort studies^31,39^. The full model equations are provided in the Supplemental Information.

**Figure 1:**
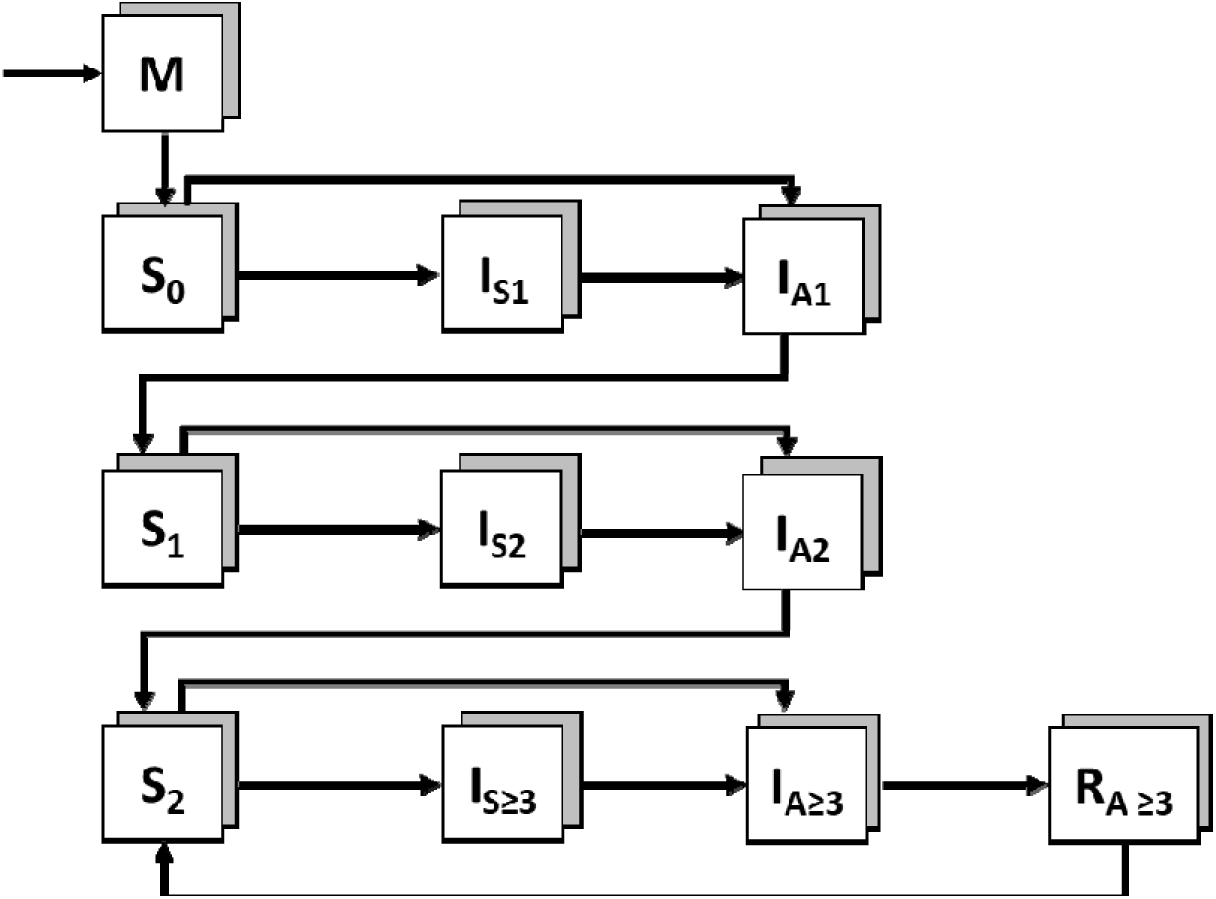
Norovirus Transmission Model Schematic: This diagram represents the model transitions and states of norovirus infection. Children are born into a maternal immunity class (M). After maternal immunity wanes, individuals move into a susceptible pool (S_0_). At the rate of the force of infection (λ_t_), individuals become infected and, based on the probability of symptomatic infection (p_S0_) move into a symptomatic (I_S1_) or directly to an asymptomatic (I_A1_) stage of norovirus infection and, after a period of symptoms, recover from symptoms but experience a period of asymptomatic infection (I_A1_). Individuals may also bypass symptomatic infection and move directly to the asymptomatic compartment (I_A1_), based on the probability of asymptomatic infection (1 - p_S0_). Following asymptomatic infection, all infected individuals recover and are again susceptible to norovirus infection (S_1_), with a reduced risk of infection (p_S1_) and symptoms (σ_1_). Norovirus immunity to infection and disease increases over three sequential infections. After the third infection (I_A≥3_), individuals enter a stage of long-term immunity (R_A≥3_). After long-term immunity is accrued, waning returns individuals to similar baseline protection as would be seen after two infections. By assuming that vaccination acts in a similar way to natural infection, we have parallel infection and immunity structure among vaccinated and unvaccinated populations, capturing increasing immune protection over a combination of three sequential vaccine doses or breakthrough infections followed by waning immunity. Under the assumption of maternal antibody interference with vaccine efficacy, children vaccinated while in the M compartment would remain in the M compartment (rather than moving up levels of immune protection). The grey and white boxes indicate the parallel vaccinated and unvaccinated model compartments or states with the lines indicating movement between model states.

#### Modeling Transmission

Individuals are born with complete protection against infection and disease (i.e., symptoms) from maternal immunity (*M*). Individuals with maternal immunity become fully susceptible (*S_0_*, numerical subscript indicating number of previous infections) to infection and disease after maternal immunity wanes (μ, rate of waning maternal immunity, Table 1). Susceptible individuals then become infected based on a seasonal force of infection, λ*(t)* (see Supplemental Materials for additional details). Infected individuals may experience asymptomatic infection (*I_A1_*) or may be symptomatic (*I_S1_*) based on a fitted symptomatic proportion (*p_S0_,* proportion symptomatic upon first infection). Symptomatic individuals progress to an asymptomatic stage of infection following their symptomatic infection. These post-symptomatic infections are grouped together with asymptomatic infections in the model and are assumed to be a less infectious (ε, multiplicative relative infectiousness during asymptomatic period) compared to symptomatic infections. Following the resolution of primary infection, individuals are again susceptible (*S_1_*) but experience a reduced force of infection (σ*_1_* < 1, where σ*_1_* is the hazard ratio for infection given 1) and reduced probability of disease given infection (*p_s1_* < 1, where *p_s1_* is the hazard ratio for symptoms given 1) for subsequent exposures. Following a secondary infection (*I_A2_, I_S2_*), individuals are again susceptible (*S_2+_*) yet experience a further reduced force of infection (σ*_2_* < σ*_1_*) and probability of disease (*p_s2_* < *p_s1_*) . Following a third infection (*I_A_*_≥_*_3_, I_S_*_≥_*_3_*), individuals are assumed to enter a maximum immunity state, with complete, temporary protection from infection (*R_3+_*). After a period of waning (θ, rate of waning of long-term immunity), individuals are again susceptible (*S_2+_*), with subsequent infections assumed to occur at the same force of infection and probability of disease as the third infection.

**Table 1:**
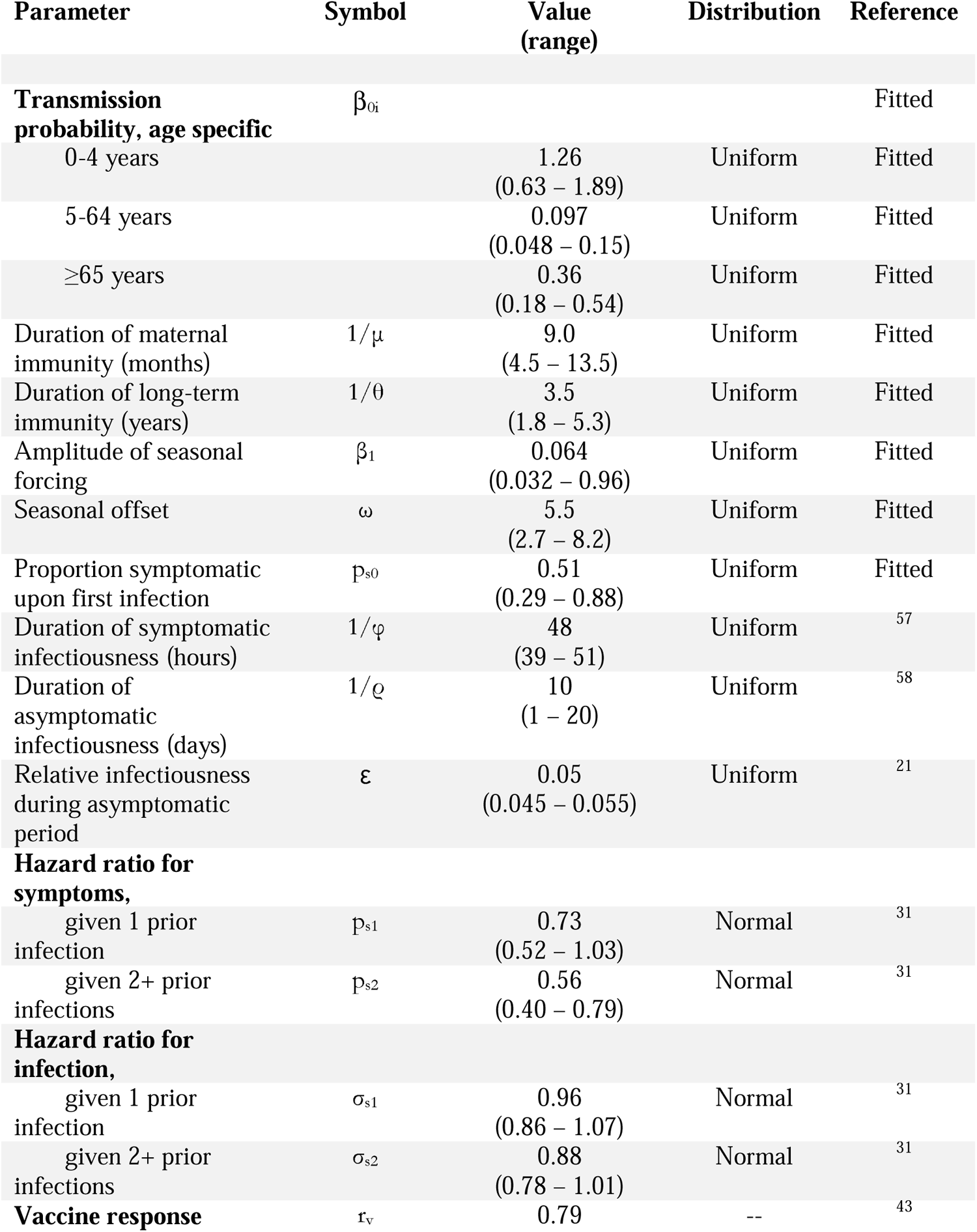
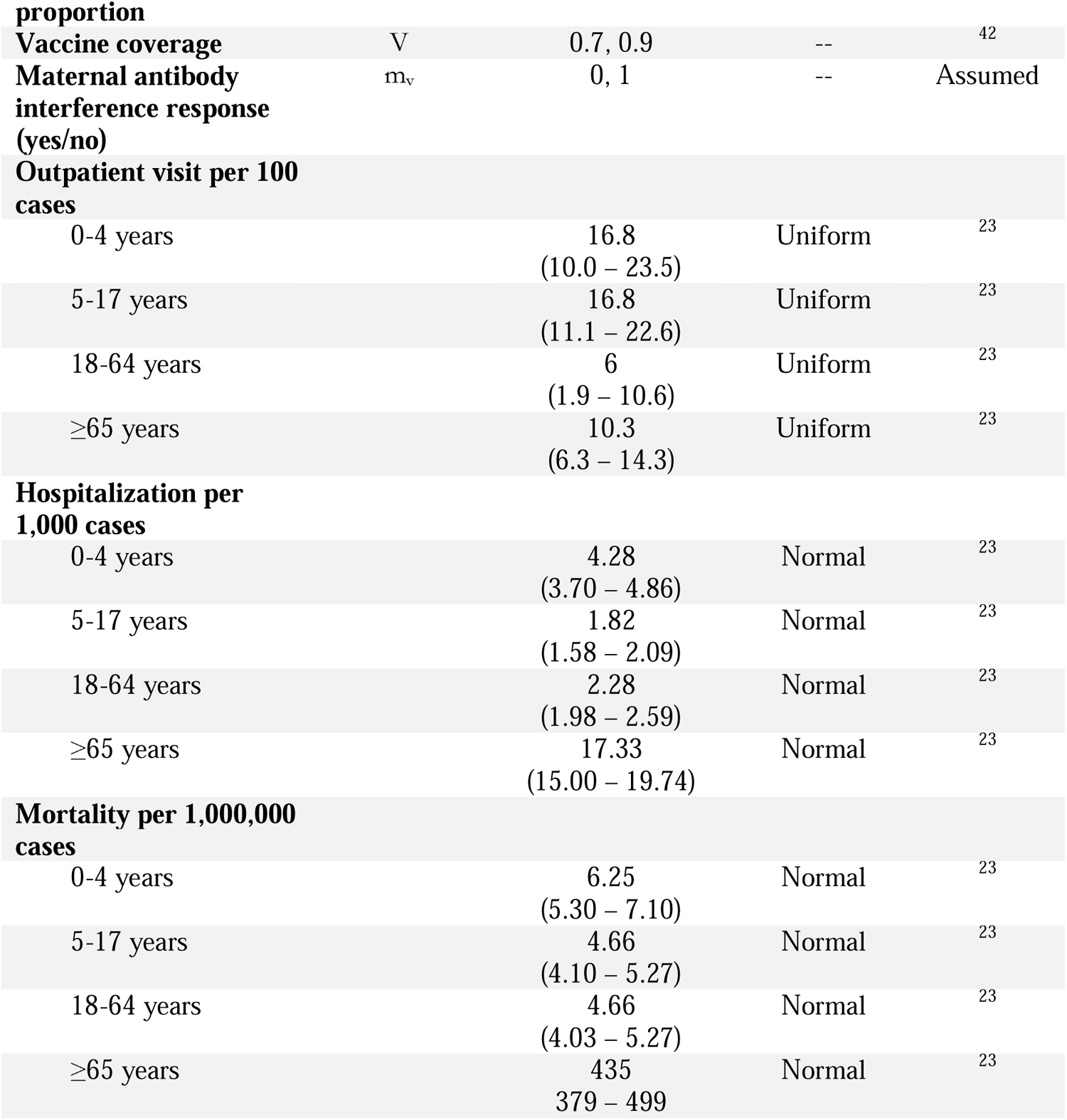
Parameter input values, ranges tested in uncertainty analyses, and source: Point values for literature-derived parameters were used for model fitting of the maximum likelihood estimate point values provided for fitted parameters. Deterministic base model outcomes are based on point values for all parameters. Value ranges and distributions are used in Latin Hypercube Sampling (LHS) for sensitivity analyses. For fitted parameters, value ranges were assumed to be ± 50% of the MLE value.

#### Modeling Vaccination

Based on the vaccination coverage (*V*), a proportion of the population receives an initial vaccine dose. We assume that all individuals who receive an initial vaccine dose complete the series. Vaccinated individuals are tracked in model compartments that parallel unvaccinated compartments. We assume that a proportion (*r*) of individuals who receive a vaccine dose do not develop an immunological response though may respond to subsequent vaccine doses. We assume that each effective vaccine dose confers the same level and duration of protection as a natural infection. Upon experiencing three exposures, through response to vaccination(s) and/or natural infection(s), individuals attain maximum immunity.

#### Age Structure

The model includes nineteen distinct age classes, with young children (<5 years of age) divided into 1-month age classes for individuals <1 year of age and 1-year age classes for children 2 to 4 years old, older children (5 - <18 years of age), adults (18 – 64 years of age), and older adults (≥65 years of age). The model tracks pediatric transmission on a finer scale within the young children age classes to allow for simulation of month-specific vaccination schedules and detailed analyses of pediatric infection, disease, and transmission. For fitting purposes, these age groups are summed into 0-6 month and 6-12 month groups to match available data. Realistic and age-specific population sizes and death rates are applied to each age group^33^, with compartment sizes initiated in each age group according to equilibrium norovirus susceptibility, infection, and immunity status. . Age groups interact based on a heterogeneous contact structure calculated from the POLYMOD study from eight European countries, which we assume to be representative of mixing patterns in the US^40^.

### Statistical Inference

We fit the model to the continuous nine-year (2011 – 2019) time series of weekly reported cases using maximum likelihood-based inference. We estimated age-specific transmission probabilities (β_0_, age groups <5 years of age, 5 – 64 years of age, ≥65 years of age), seasonality coefficients (β_1_, ω), rate of waning of maternal immunity (μ), rate of waning of long-term immunity (θ), and proportion symptomatic upon initial infection (p_s0_). To obtain the maximum likelihood estimates (MLE), we calculated individual negative log-likelihoods (NLL) for each age group, assuming the reported weekly cases scaled to the US population, were Poisson distributed with the mean and variance equal to the model-predicted weekly cases multiplied by the age-specific reporting rate (see Supplementary Material). We summed individual NLLs to calculate an overall NLL and estimated the best-fit parameters by minimizing the NLL for all age groups combined.

### Model Scenarios

#### Vaccination Schedules

We simulated five vaccination schedules: (1) three dose series administered at 0, 2, and 4 m of age (0/2/4m); (2) three dose series administered at 2, 4, and 6 m of age (2/4/6m); (3) two dose series administered at 6 and 12m of age (6/12m); (4) two dose series administered at 12 and 24 m of age (12/24m); and (5) one dose administered at 48 m of age (48m). These schedules represent vaccination time points corresponding with existing ACIP-recommended pediatric vaccination schedules^41^. Vaccination ages (excluding birth) and number of doses are also consistent with the Phase II norovirus vaccine trial that included children from 6 weeks to 4 years of age^17^. The birth dose, which is outside of the age range of Phase II trials, was included as a comparison with previous norovirus transmission models, which implicitly simulated vaccination at birth^22,38^.

#### Vaccine Coverage and Characteristics

For each vaccination scenario, we simulated lower (70%) and higher (90%) coverage (V). Coverage levels were selected based on the range of vaccination coverage for currently recommended pediatric vaccinations in the US: rotavirus vaccination coverage at approximately 70%, MMR and DTaP vaccination coverage at ≥90% for a complete series of doses^42^. Higher (90%) coverage results are included in the Supplementary Materials.

We calculated the per dose vaccine response probability (*r*) based on the two-dose seroresponse proportion for children 6 months to <1 year of age from the Phase II safety and immunogenicity trial conducted for the Takeda bivalent VLP vaccine in children, 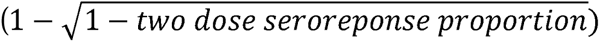, assuming vaccine response for the two doses is independent^43^. This dose response probability was applied to each vaccine dose for all vaccination schedules, with exceptions for maternal antibody inference with vaccination described below. We assumed that a dose that elicited an immune response conferred protection equivalent to one natural infection and an ineffective dose did not confer any additional protection.

#### Maternal Antibody Interference

We considered the impact of maternal antibody interference on overall vaccine impact for all vaccination schedules. For the no maternal antibody interference scenarios (*m_v_* = 1), we assumed that individuals vaccinated while in the maternal immunity (M) compartment would respond to vaccination in the same manner as other individuals. For the maternal antibody interference scenarios (*m_v_*= 0), we assumed complete interference with vaccination, such that individuals vaccinated while in the M compartment do not respond to vaccination and receive no additional protection from vaccination. To simulate complete maternal antibody interference, we set the dose response probability *r* to 0 for individuals vaccinated while in the M compartment. In reality, maternal norovirus antibodies are likely to partially interfere with vaccine take, similar to observations for rotavirus vaccination and maternal rotavirus antibodies (transferred both from placenta and breast milk)^44,45^. As such, these scenarios are meant to represent the possible bounds of the influence of maternal immunity and antibody interference on overall vaccine impact.

### Outcomes

We compared norovirus AGE outcomes with and without vaccination for each vaccination schedule, coverage level, and maternal antibody interference scenario. For each scenario, we calculated the percent norovirus AGE cases averted annually and cumulatively by age group and for the total population over five years following the introduction of vaccination. Additionally, we investigated the indirect effects of vaccination by comparing the difference between the incidence of norovirus AGE cases predicted with vaccination and a dynamic force of infection with the incidence of norovirus AGE cases predicted with no vaccination and a fixed force of infection^35^. We estimated vaccination impact on outpatient visits, hospitalization, and death by multiplying the predicted age-specific norovirus AGE incidence by age-specific outcome probabilities^23^.

To assess the influence of uncertainty in model parameters, we estimated the range of outcomes in a probabilistic sensitivity analysis using Latin Hypercube Sampling (LHS) of parameter ranges and distributions (Table 1). For MLE parameters, LHS was conducted over a range of ±50% of the MLE value. Using 1000 parameter draws, we estimated the 2.5^th^ and 97.5^th^ percentile uncertainty intervals (UI) and the partial rank correlation coefficients for annual percent cases averted at five years after vaccination introduction among young children (<5 years) and the total population. In this way, we captured the range of possible outcomes generated by uncertain parameters. Additional details are provided in the Supplemental Materials.

Model fitting, simulation, and analyses were conducted in R (version 4.0.5), utilizing the nloptr package for model fitting and the deSolve package and EpiModel^46^ package for model implementation and simulations.

## Results

### Model fit

Predicted cases by the MLE-parameterized model capitulated key features of the observed data, including seasonal trends as well as age-specific and total annual reported cases (Figure 2). Adjusting for under-reporting, the MLE model estimated a total annual incidence of 19.7 million AGE cases, consistent with the 19-21 million AGE cases attributed to norovirus annually in the United States^8^. MLE parameters for the model were also biologically reasonable, with the duration of maternal immunity estimated at 9.0 months and the duration of long-term immunity estimated at 3.5 years (Table 1). Additional details on alternative fits are included in the Supplemental Materials.

**Figure 2:**
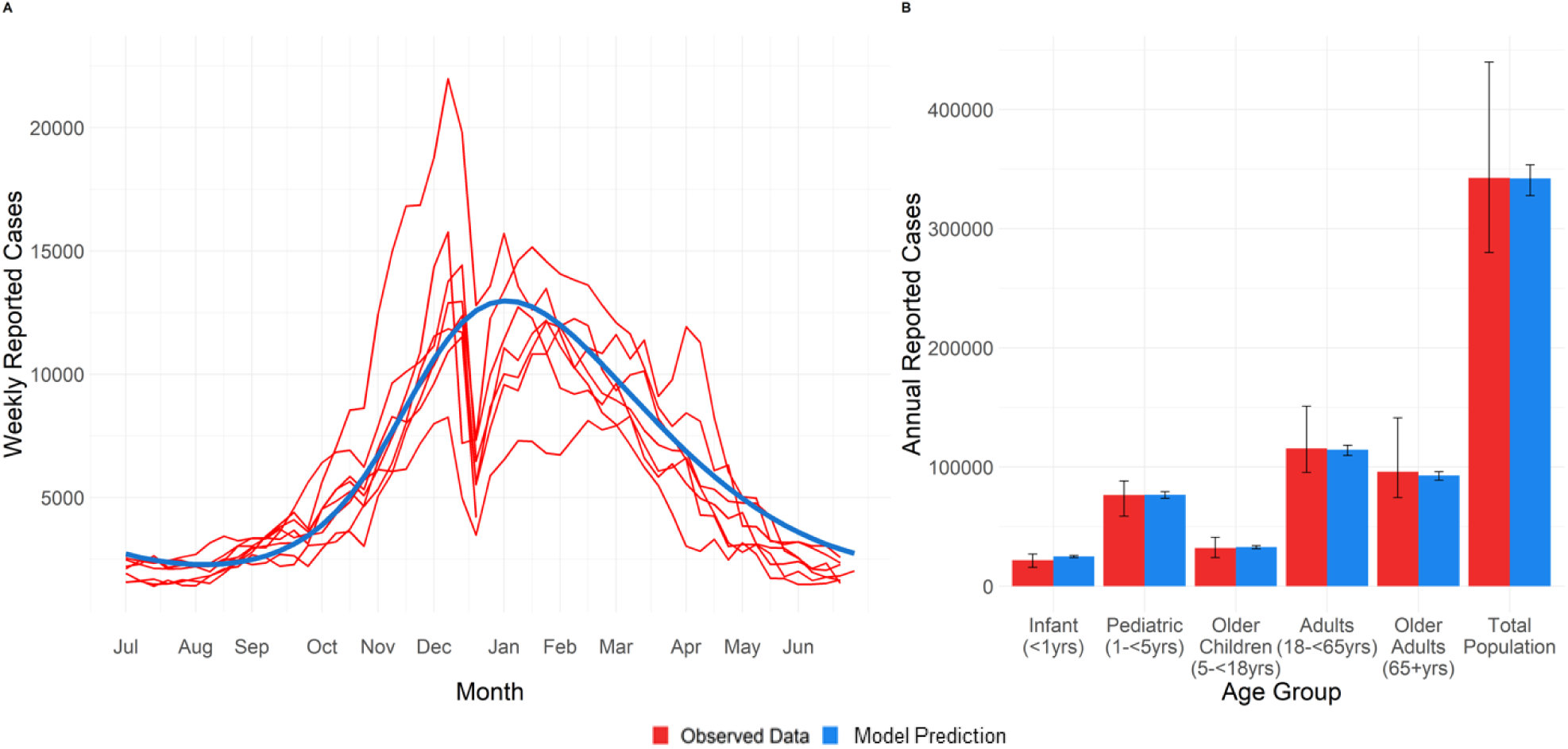
Observed (red) and MLE model-predicted (blue) reported norovirus cases without vaccination. (a) total weekly cases (each red line represents one year of data, 2011 – 2019, the blue line represents the steady-state fitted model data; fit for age group-specific weekly cases presented in the Supplemental Materials), (b) average annual cases by age group (error bars represent the of annual reported norovirus cases over the nine year period).

### Vaccine impact

#### Overall Vaccination Impact

Vaccine impact is described as the annual rate of cases averted among young children (cases averted per 100,000 children <5 years of age) and cases averted across the total population (cases averted per 100,000 people). Over the five years following the introduction of vaccination, the impact of vaccination increased among young children and the total population before stabilizing. The greatest stabilized vaccine impact was 12,500 cases averted annually per 100,000 young children and 1,200 cases averted annually per 100,000 people across the total population, given 70% vaccination coverage and no maternal antibody interference with vaccine efficacy (Figure 3A). The reduction of cases due to vaccination was greatest during the summer and fall, outside the traditional norovirus season, and was reduced in the later winter and early spring, during the norovirus season (Figure 3B). This seasonal pattern became more pronounced as vaccination became more established in the population and was similar regardless of maternal antibody interference scenario and vaccination coverage level (Figure 3 for no maternal antibody interference and 70% vaccination coverage, SI Figure 2 for complete antibody interference and 70% and SI Figures 3 and 4 for 90% vaccination coverage with no maternal antibody interference and with complete maternal antibody interference).

**Figure 3:**
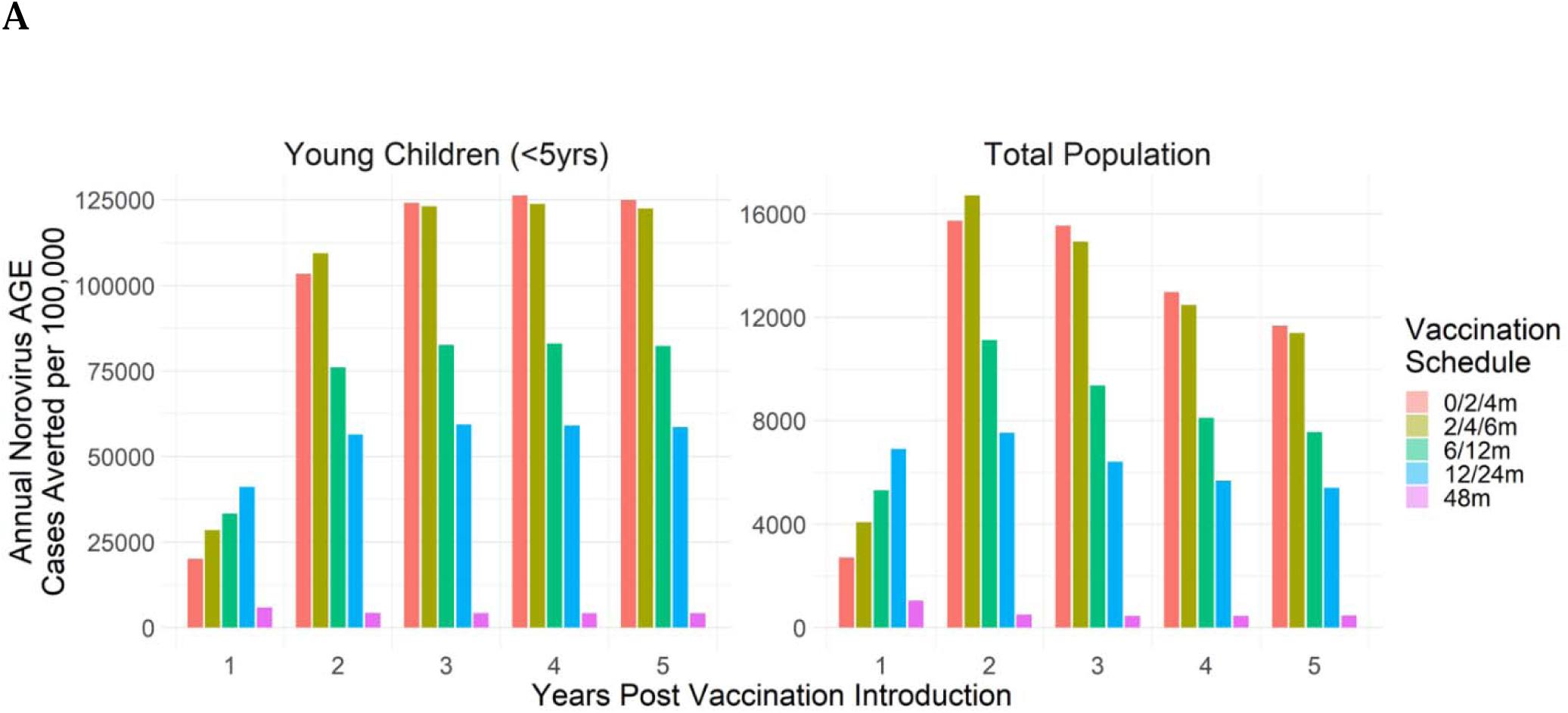
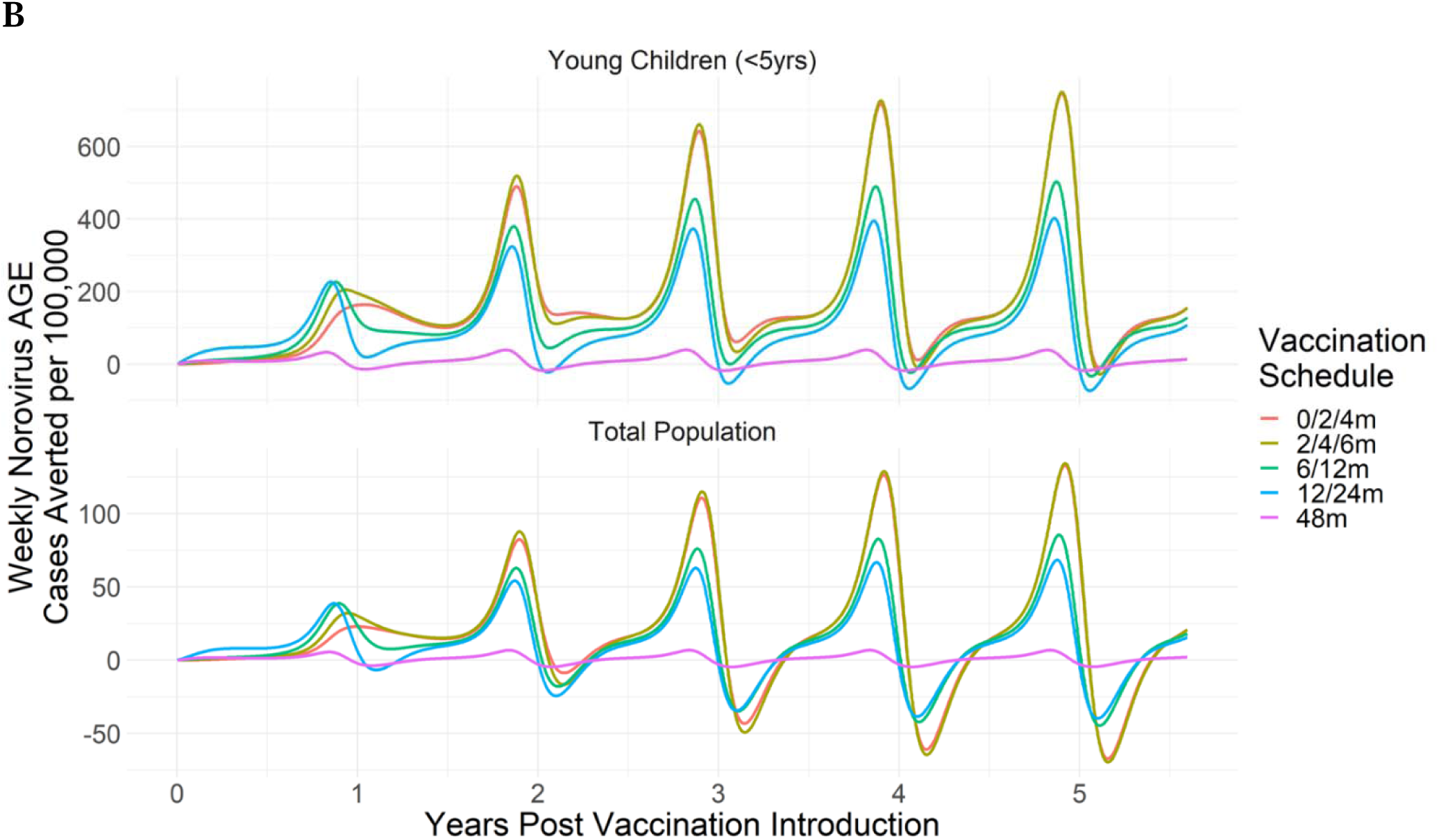
Norovirus AGE cases averted for five years following the introduction of vaccination as (a) annual cases averted per 100,000 and (b) weekly cases averted per 100,000, given 70% vaccination coverage and no maternal antibody interference for young children (<5 years of age) and total population across vaccination schedules. Note that incidence rates are per either 100.000 young children or 100,000 total population.

#### Impact by Vaccine Schedule

In the first year following vaccine introduction, assuming no maternal antibody interference and 70% vaccination coverage, the two-dose 12/24m vaccination schedule was most impactful and cases averted decreased with earlier vaccination schedules (Figure 3A). By five years post vaccine introduction, the three-dose 0/2/4m and 2/4/6m schedules were established as the schedules with the greatest impact, compared to other schedules. The 0/2/4m schedule averted 44% (95% UI: 30 – 64%) of cases in young children, between 6% (95% UI: 1– 54%) and 11% (95% UI: 2 – 58%) of cases in older age groups, and 19% (95% UI: 7 – 60%) of cases in the total population, with very similar impacts for the 2/4/6m vaccination schedule. Across age groups, later vaccination, compared to earlier vaccination, resulted in decreased impact, with the 48m schedule having the least impact. The 48m schedule was the least impactful in young children (2% [95% UI: 1 – 6%] cases averted) and overall (1% [95% UI: 0 – 5%] cases averted) by a substantial margin and was associated with a slight increase in cases among older children (-2% [95% UI: -19 – 1%] cases averted).

With 90% vaccination coverage, at five years post vaccine introduction, the most impactful pediatric norovirus vaccination schedules remained the 0/2/4m and the 2/4/6m schedules, averting 55% (95% UI: 39 – 92%) and 54% (95% UI: 38 – 91%) of norovirus AGE cases among young children and 25% (95% UI: 9% - 90%) and 25% (9% - 89%) of cases across the total population, respectively (SI Figure 5). Across all simulated vaccination schedules, increasing vaccination coverage by 20%, from 70% to 90%, increased the number of cases averted by 27% (48m schedule) to 33% (12/24m schedule) in the total population. While a greater impact was demonstrated with 90% vaccination coverage compared to 70% coverage, the patterns by vaccination scenario and years post vaccination introduction were similar (SI Figure 5, Figure 3A).

#### Impact of Maternal Antibody Interference

Among young children, the effect of maternal antibody interference reduced vaccine impact by 82% for the 0/2/4m schedule (maximum reduction observed), and by 68% for the 2/4/6m schedule (Figure 4). For vaccine schedules beginning at 6m or later, the effect of maternal antibody interference was substantially lessened, with a 46% reduction in vaccine impact associated with maternal antibody interference for the 6/12m schedule, a 31% reduction for the 12/24m schedule, and virtually no difference by the 48m schedule. When assuming maternal antibody interference, given 70% vaccination coverage, the 6/12m vaccination schedule was most impactful, averting 16% (95% UI: 7 – 25%) of norovirus AGE cases among young children and 7% (95% UI: 2 – 24%) of cases across the total population with similar impacts produced by vaccination under the 2/4/6m and 12/24m schedules (Figure 4, SI Table 1). Considering the impact of vaccination with and without maternal antibody interference, vaccination at 2/4/6m was consistently among the most impactful schedules, while the relative impact of other schedules was more dependent on maternal antibody interference assumptions.

**Figure 4:**
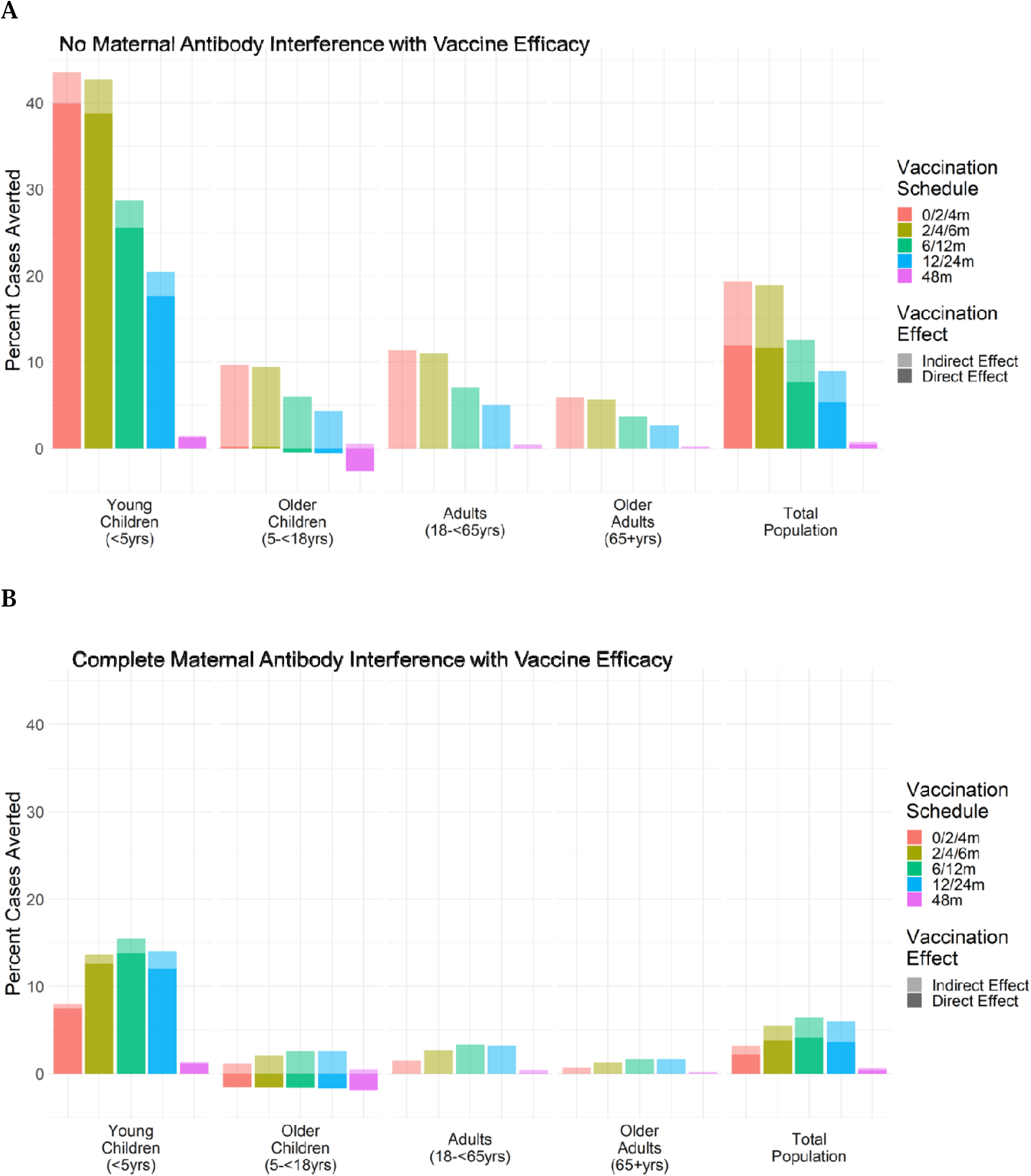
Annual direct and indirect effects of pediatric norovirus vaccination as percent norovirus AGE cases averted, (a) with no maternal antibody interference with vaccine efficacy and (b) with complete maternal antibody interference with vaccine efficacy : Different vaccine schedules are indicated by bar colors. Direct vs indirect effects represented by dark vs light opacity. Each plot represents the annual percent cases averted for a single year after five years of vaccination. Percent cases averted are presented for individual age groups and the total population.

#### Indirect Effects

Approximately 67% of the reduction in disease from pediatric vaccination occurred among young children, in the vaccinated group (Figure 4). However, a substantial proportion of vaccination impacts occurred in nonvaccinated individuals, with 20% of all cases averted occurring among adults, a nonvaccinated age group, reflecting the indirect effects of norovirus vaccination resulting from decreased norovirus circulation in this age group.

To capture the total impact of indirect effects in all age groups, we fixed the force of infection to its prevaccine value, as described in the methods. With no maternal antibody interference, 4% (95% UI: 2 – 7%) of norovirus AGE cases were averted in young children by indirect effects (70% coverage, 0/2/4m, 2/4/6m schedules, Figure 4A), representing 9% of the overall impact of vaccination on norovirus cases in young children. Across the total population, 7% (95% UI: 4 – 14%) of norovirus AGE cases were averted by indirect effects, representing 38% of the overall impact of vaccination on norovirus cases across the population. With maternal antibody interference, the relative indirect effect was dampened. For the most impactful vaccination schedule with maternal antibody interference (6/12m), indirect effects were associated with averting 2% (95% UI: 1 – 3%) of norovirus AGE cases among young children (13% of the overall impact of vaccination on norovirus cases in young children) and 2% (95% UI: 1 – 6%) of total norovirus AGE cases in the total population (36% of the overall impact of vaccination on norovirus cases across the population, Figure 4B).

#### Clinical Outcome Impacts

Expanding from relative changes in norovirus AGE cases to total outcomes averted across clinical outcomes, among young children, norovirus vaccination at five years was estimated to annually avert up to 12,500 AGE cases (95% UI: 7,200 – 22,200), 2,100 outpatient visits (95% UI: 900 – 4,600), 50 hospitalizations (95% UI: 30 – 100), and 0.1 deaths (95% UI: 0.04 – 0.14) per 100,000 young children (Table 2, assuming no maternal antibody interference, 70% vaccination coverage, 0/2/4m schedule). In the total population, norovirus vaccination annually averted up to 11,700 AGE cases (95% UI: 6,500 – 22,000), 1,600 outpatient visits (95% UI: 800 – 3,400), 60 hospitalizations (95% UI: 30 – 130), and 0.5 deaths (95% UI: 0.16 – 1.91) per 1 million people. In total counts of outcomes averted, this equates to norovirus vaccination averting annually 2.6 million AGE cases (95% UI: 1.48 – 4.55 million), 431,000 outpatient visits (95% UI: 190,000 – 942,000), 11,000 hospitalizations (95% UI: 56,300 – 19,800), and 16 deaths (95% UI: 9 – 29) among young children and 3.8 million AGE cases (95% UI: 2.14 – 7.21 million), 539,000 outpatient visits (95% UI: 254,000 – 1,117,000), 18,500 hospitalizations (95% UI: 9,800 – 43,600), and 158 deaths (95% UI: 52 – 623) across the total population. A smaller proportion of severe outcomes were averted among the total population (7% of total deaths averted) compared to less severe outcomes (24% of total outpatient visits averted).

**Table 2:**
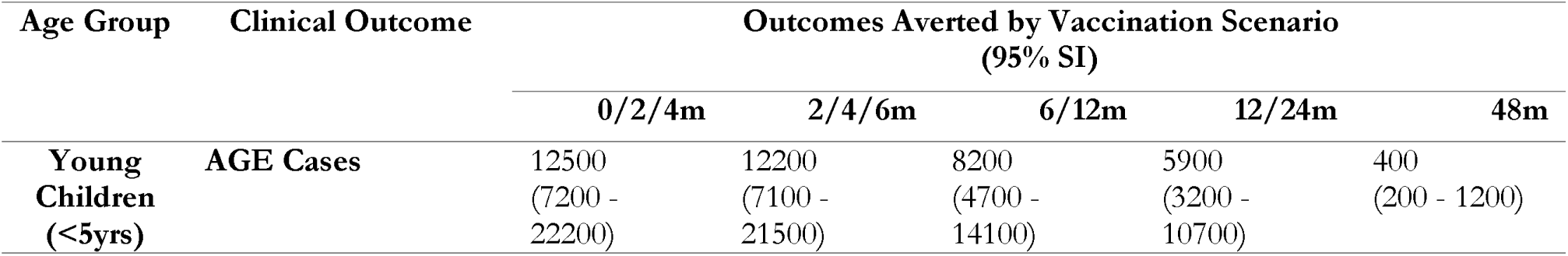
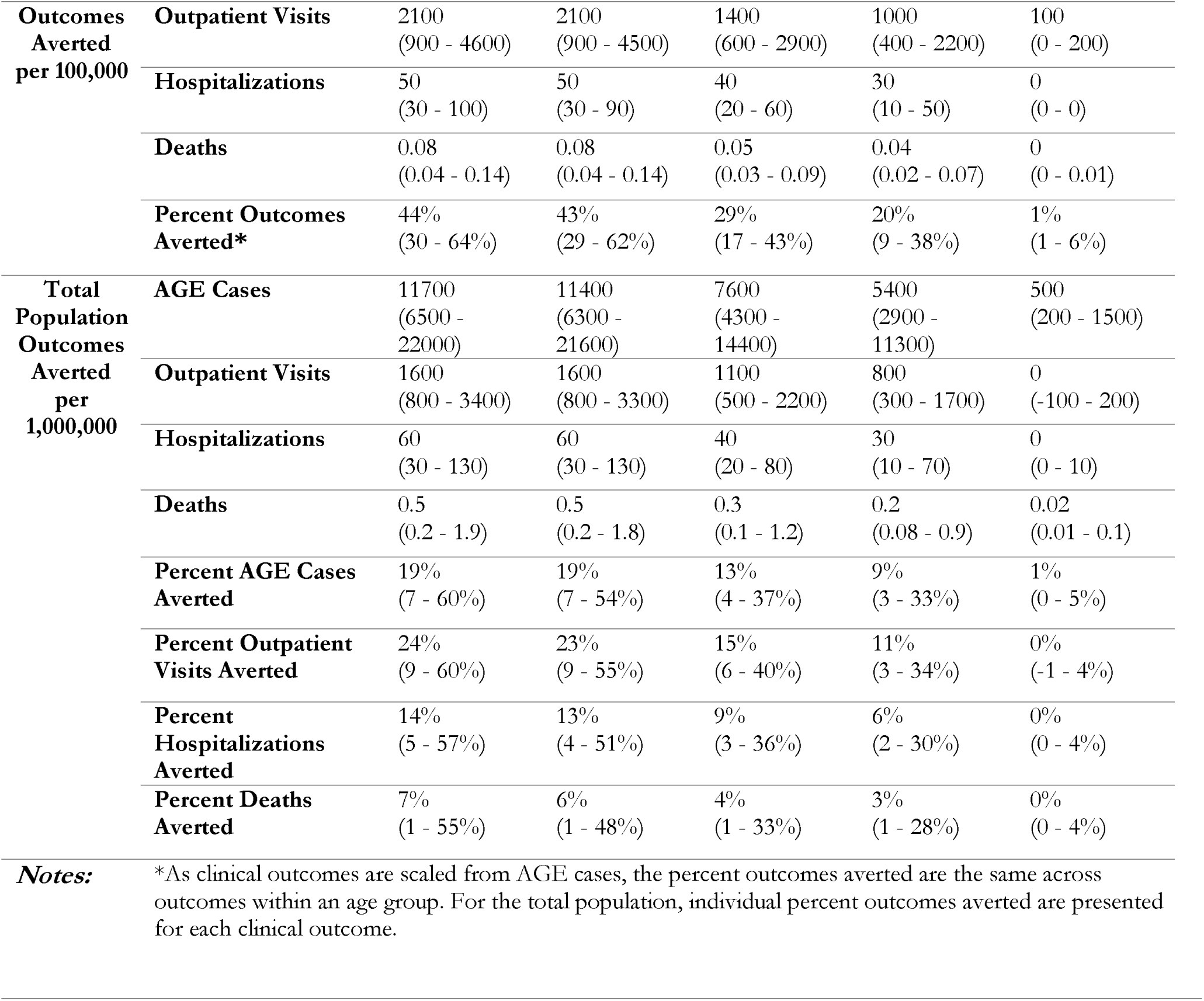
Total clinical outcomes averted annually by vaccination scenario (95% uncertainty interval based on LHS sensitivity analysis) by age group. Results associated with no maternal antibody interference with vaccine efficacy and 70% vaccination coverage.

#### Vaccine Efficiency

Over the five years following the introduction of vaccination, without maternal antibody interference, vaccination cumulatively averted up to 0.5 cases per dose (95% UI: 0.3 – 0.8, 6/12m schedule) and 0.13 cases per vaccinee (95% UI: 0.7 – 2.1, 0/2/4m and 2/4/6m schedule) in the total population (Table 3). This equates to 2.2 doses administered (or 0.8 individuals fully vaccinated) to avert a single case. With maternal antibody interference, vaccine impact and, subsequently, vaccine efficiency was lower (SI Table 2).

**Table 3:** Vaccine efficiency by vaccination scenario (95% uncertainty interval based on LHS sensitivity analysis) by age group. Results associated with no maternal antibody interference with vaccine efficacy and 70% vaccination coverage. Vaccine efficiency measures are presented for the cumulative doses or vaccinated individuals and cumulative cases averted over five years.

| *Efficiency Measure* | *Efficiency by Vaccination Scenario (95% UI)* |  |  |  |  |
| --- | --- | --- | --- | --- | --- |
|  | **0/2/4m** | **2/4/6m** | **6/12m** | **12/24m** | **48m** |
| Doses to avert a case | 2.3<br>(1.4 - 4.3) | 2.3<br>(1.3 - 4.2) | 2.2<br>(1.2 - 4) | 2.9<br>(1.4 - 5.3) | 16.7<br>(5.2 - 36) |
| Vaccinees to avert a case | 0.8<br>(0.5 - 1.5) | 0.8<br>(0.5 - 1.5) | 1.1<br>(0.7 - 2.1) | 1.5<br>(0.8 - 2.9) | 16.7<br>(5.2 - 36) |
| Cases averted per dose | 0.4<br>(0.2 - 0.7) | 0.4<br>(0.2 - 0.7) | 0.5<br>(0.3 - 0.8) | 0.3<br>(0.2 - 0.7) | 0.1<br>(0 - 0.2) |
| Cases averted per vaccinee | 1.3<br>(0.7 - 2.1) | 1.3<br>(0.7 - 2.1) | 0.9<br>(0.5 - 1.5) | 0.7<br>(0.3 - 1.2) | 0.1<br>(0 - 0.2) |

## Discussion

We aimed to compare the impact of five different pediatric norovirus immunization schedules aligned with current US ACIP-recommended visits. We found that, vaccination at 2/4/6m is likely to be the most impactful pediatric norovirus vaccination schedule across assumptions, reducing norovirus burden among young children by up to 43% assuming 70% vaccination coverage. Additionally, pediatric vaccination at 2/4/6m averted 11,400 cases, 1,600 outpatient visits, 60 hospitalizations, and 0.5 deaths per 1 million people, or a total of 3.72 million cases, 527,000 outpatient visits, 18,000 hospital visits, and 152 deaths across the total population, with 70% vaccination coverage. Our results show that pediatric vaccination has the potential to avert a substantial number of episodes for older populations. Taken together, these results suggest that even when the complexities of early norovirus infection and immunity dynamics are considered, implementing pediatric norovirus vaccination would have a substantial impact on pediatric and overall norovirus burden.

### Most impactful schedules and maternal antibody interference assumptions

While vaccination at 2/4/6m was not the most optimal schedule in either scenario (i.e., with or without maternal antibody interference), this schedule most effectively balanced tradeoffs between (a) vaccinating early enough to protect children during the period of greatest risk and (b) vaccinating late enough to not be overly impacted by maternal antibody interference. Without maternal antibody interference, the 0/2/4m and 2/4/6m schedules were similarly impactful; with interference, the 0/2/4m was substantially less so. The period of greatest risk is between 6 to 24 months of age, with lower norovirus AGE incidence prior to 6 months of age^4,29,39^. Since norovirus incidence is low in the first 2 months of life, there is little benefit to starting the vaccine schedule at birth instead of at 2 months. While later vaccination was more impactful in the presence of maternal antibody interference, the 2/4/6m schedule fell only slightly behind the 6/12m and 12/24m schedules in terms of greatest impact under this assumption. The similarity between the impacts of these schedules reflects the mechanism of modeling maternal antibody waning. In our model, we assumed exponential waning of maternal immunity, with the best fit duration of maternal immunity of 9 months such that approximately 20% of children by 2 months and over 50% of children by 6 months have lost maternal protection. This mechanism of exponential waning also means that over half of vaccinated children receive an effective vaccine dose under the 2/4/6m schedule even with maternal antibody interference. These estimates of waning duration are consistent with immunological studies, which show a decrease in norovirus immunoglobulin G titers from birth to six months of age^24^.

For simplicity, we assumed maternal antibodies interference was ‘all-or-nothing’. In reality, maternal antibodies likely provide partial protection from norovirus infection and partially interfere with vaccine immunogenicity, as suggested by infection and serology data for norovirus and other pathogens including rotavirus^24,28,29,45,47^. While these studies provide some evidence of maternal antibody interference, they also reveal the limited understanding of the impact of maternal antibodies and maternal immunity on norovirus infection and disease in young children.

The ability of vaccination to protect children throughout the period of greatest norovirus risk also relates to the waning of vaccine and natural immunity. Our estimate of duration of immunity of 3.5 years falls between estimates from challenge (6 months to 2 years) and modeling studies (4.1 – 8.7 years), with this wide range representing uncertainty regarding norovirus immunity^21,30,48,49^.

In our sensitivity analysis, we found that specific model results were strongly affected by the duration of maternal (SI Figure 7), and natural immunity, such that longer duration of immunity was associated with greater vaccination impact. (SI Figures 6A/6B). Across all immunity sensitivity analyses, we found that the relative impacts of pediatric vaccination schedules were robust and the 2/4/6m schedule remained among the most impactful (SI Figures 5A/5B)

Still, further investigation of norovirus maternal immunity and potential interference with vaccination is warranted to better understand the potential population impacts of pediatric norovirus vaccination. This may be particularly important to consider in low and middle-income countries, where norovirus incidence is higher^1,26^.

### Indirect Effects and Severe Clinical Outcomes

Consistent with findings from previous modeling studies, we found that pediatric vaccination was able to produce indirect effects across the population^22,38,50,51^. However, indirect effects among older adults (65+ years of age) were modest compared to previous studies, which found the greatest indirect impacts among this age group^22,50,51^. These results indicate potential limits in using pediatric vaccination to avert severe outcomes. While the majority of medically-attended norovirus cases are experienced by children, older adults experience higher rates of severe clinical outcomes (i.e., hospitalization and death)^8,15,20^. With just 6% of cases averted among older adults, it may be necessary to offer vaccination to populations at risk for severe outcomes, such as older adults and particularly those living in long term care facilities, to reduce severe outcomes more substantially.

Overall, we found more limited indirect effects compared to prior models^22,38,50,51^. This may be a result of the complex norovirus infection and immunity structure, within our model compared to other models or types of infections^22,38^, particularly the restructuring of symptomatic versus asymptomatic infection allowing for asymptomatic infection alone.

We found that indirect effects were greatest among younger adults (18 – 65 years of age), differing from prior studies^22,50,51^. This may highlight the potential role that children play in transmission of norovirus to adult caretakers^21^ and the indirect impacts that infections in young children have on further transmission among adults. By including smaller age classes among young children, our model may have been more equipped to capture this dynamic. Our model also produced negative percent cases averted among older children (5 - <18 years of age), particularly when vaccinating at 48m, which may indicate shifting in age of infection from young children to older children as vaccination pushes off infections that may have occurred earlier in life but now occur later, after vaccine-derived immunity has waned.

### Limitations

We made several simplifying assumptions in this model. We did not consider viral diversity, effectively assuming equal protection across strains with natural and vaccine-induced immunity. In reality, noroviruses are a genetically diverse group of RNA viruses, with limited cross-protection across genogroups^52^ and can rapidly evolve with the potential to evade host immunity^48,53,54^. Our model parameters, most relevantly the duration of long-term immunity, were estimated from case data which included a diversity of infecting genogroups and encompassed years in which the predominant norovirus strain changed^55,56^. However, vaccine strategies may need to be adapted depending on evolving norovirus strains. Analysis of heterotypic natural and vaccine immunity and genetic differences in host response to norovirus infection could inform these strategies. Additionally, we assumed homogeneous contact structures within age groups. In reality, children who attend day care, parents, childcare providers, among others, may have contacts which allow for higher rates of norovirus transmission. Including heterogeneous contact structures, particularly greater contacts with children, may impact indirect effects of pediatric vaccination among these populations. Lastly, we assumed that norovirus vaccination provides equivalent immune protection to natural infection. Most likely, vaccination is less effective than natural infection, which may shift dynamics of the potential impact of vaccination and influence vaccination strategies.

### Conclusion

In conclusion, across a range of vaccination schedules and maternal antibody interference assumptions, norovirus vaccination at 2/4/6m consistently was found to be an impactful strategy, particularly in pediatric populations. The 2/4/6m schedule aligns with several other pediatric vaccination schedules, including diphtheria, tetanus and acellular pertussis (DTaP), pneumococcal conjugate (PCV13), inactivated poliovirus (IPV), and rotavirus in the current US ACIP-recommended vaccination schedule^41^. Integrating with the current schedule could facilitate vaccine uptake, yet also raises the concern of adding another vaccination to a crowded vaccination schedule. Incorporating trial-based vaccine response data along with coverage levels informed by the National Immunization Survey and schedules built around ACIP recommendations, allowed us to increase the realism of the transmission model and vaccination scenarios. Trial data on vaccine efficacy in children and studies on vaccine willingness can refine our models. Additionally, models expanded to consider multiple norovirus strains and cross-protection will enhance realism. Finally, models should be adapted to low- and middle-income settings to tailor recommendations to these settings, where pediatric norovirus incidence is higher and disease burden is greater^1,26^.

## Data Availability

All data and code produced in the present study are available upon reasonable request to the authors.

