## Supplemental Information for "Pediatric Norovirus Vaccination: Modeling the Influence of Schedule on Population Health"

**Supplemental Materials**

**Supplemental Text**

1. **Detailed model description**

The ordinary differential equations constituting the norovirus transmission model as shown in Figure 1 are as follows:

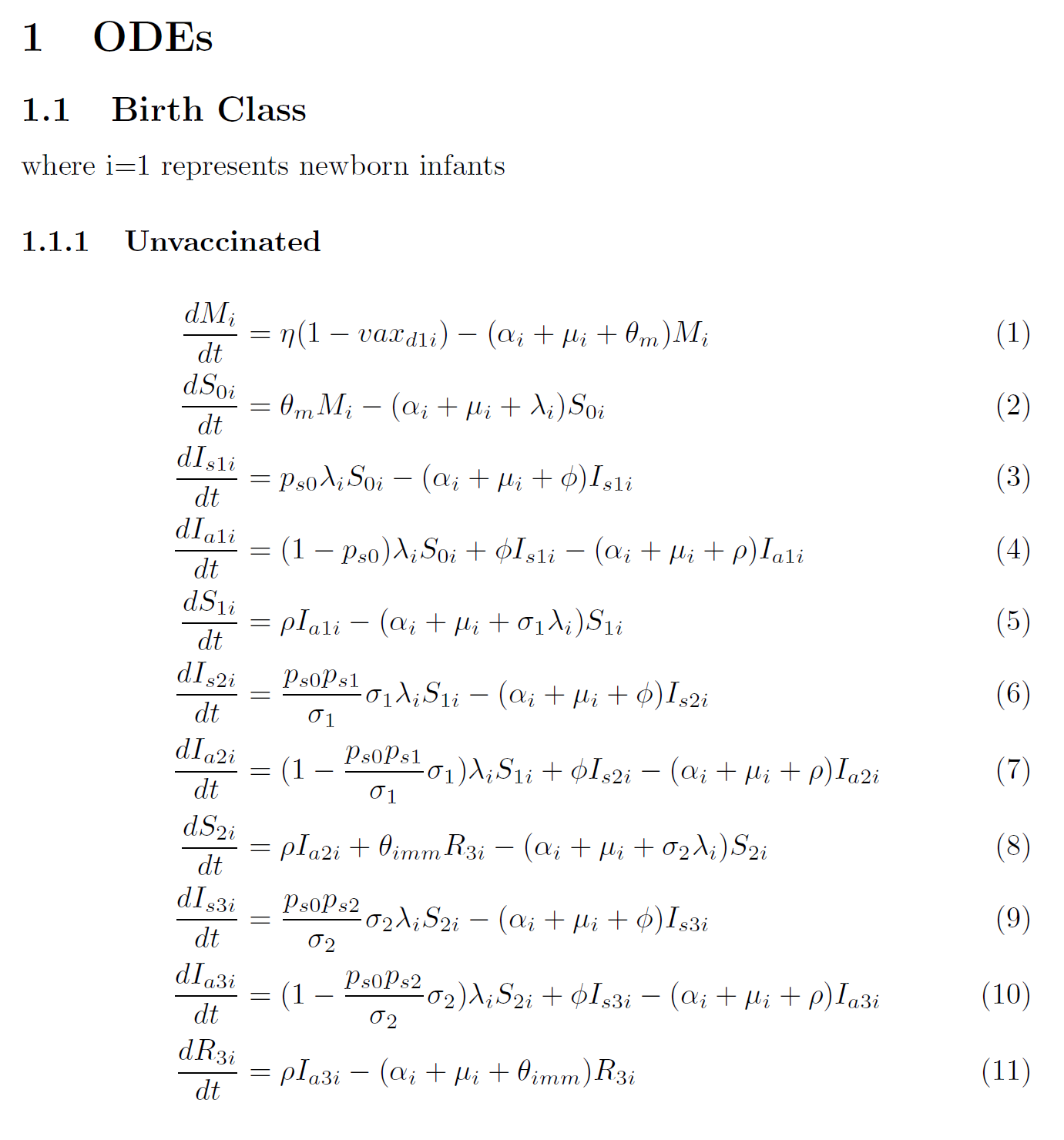

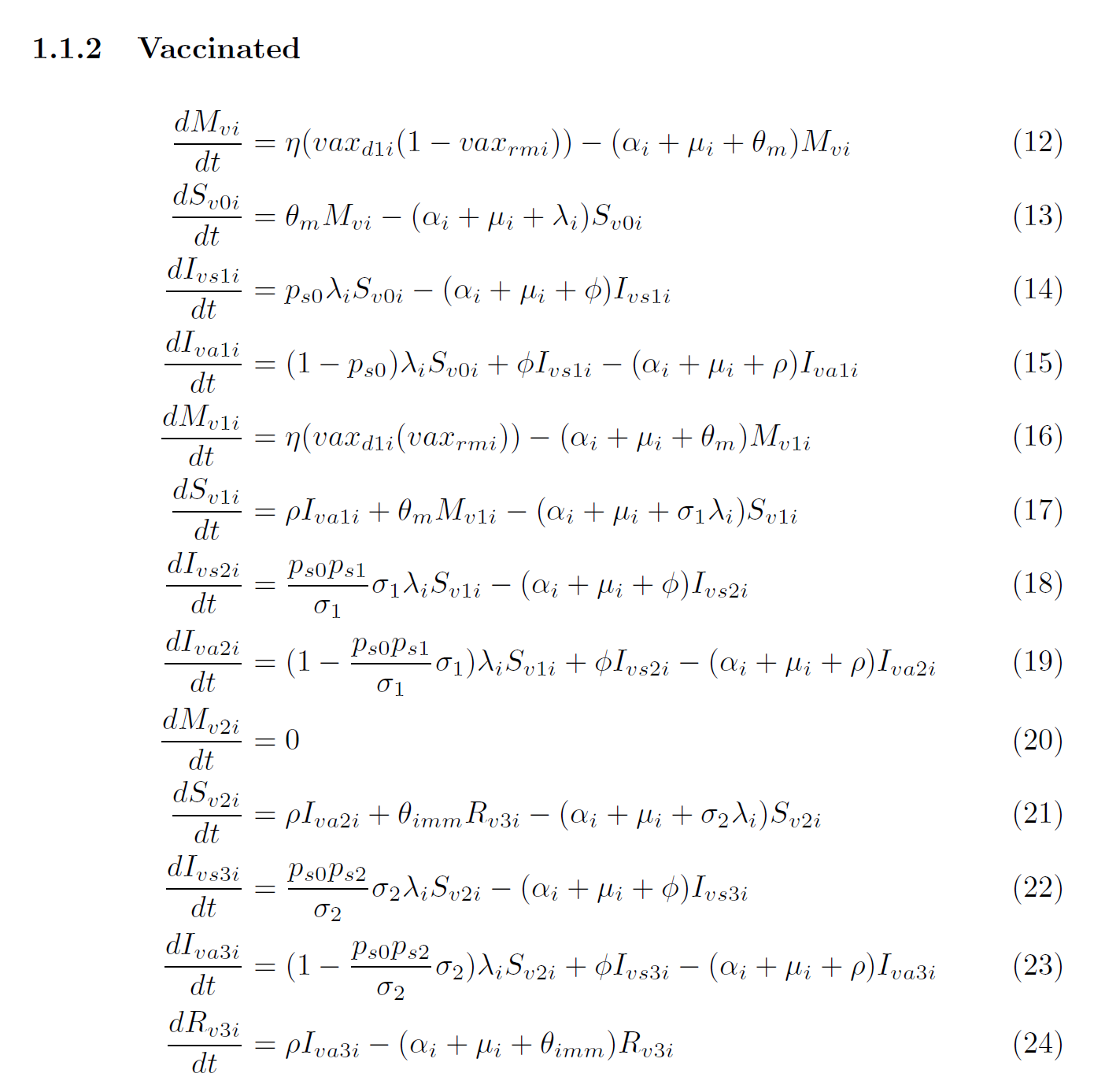

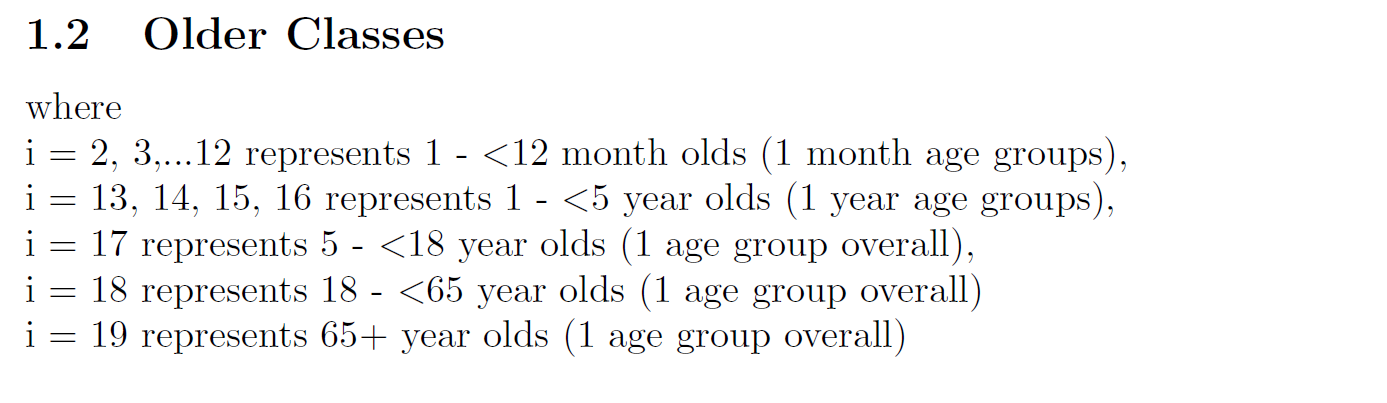

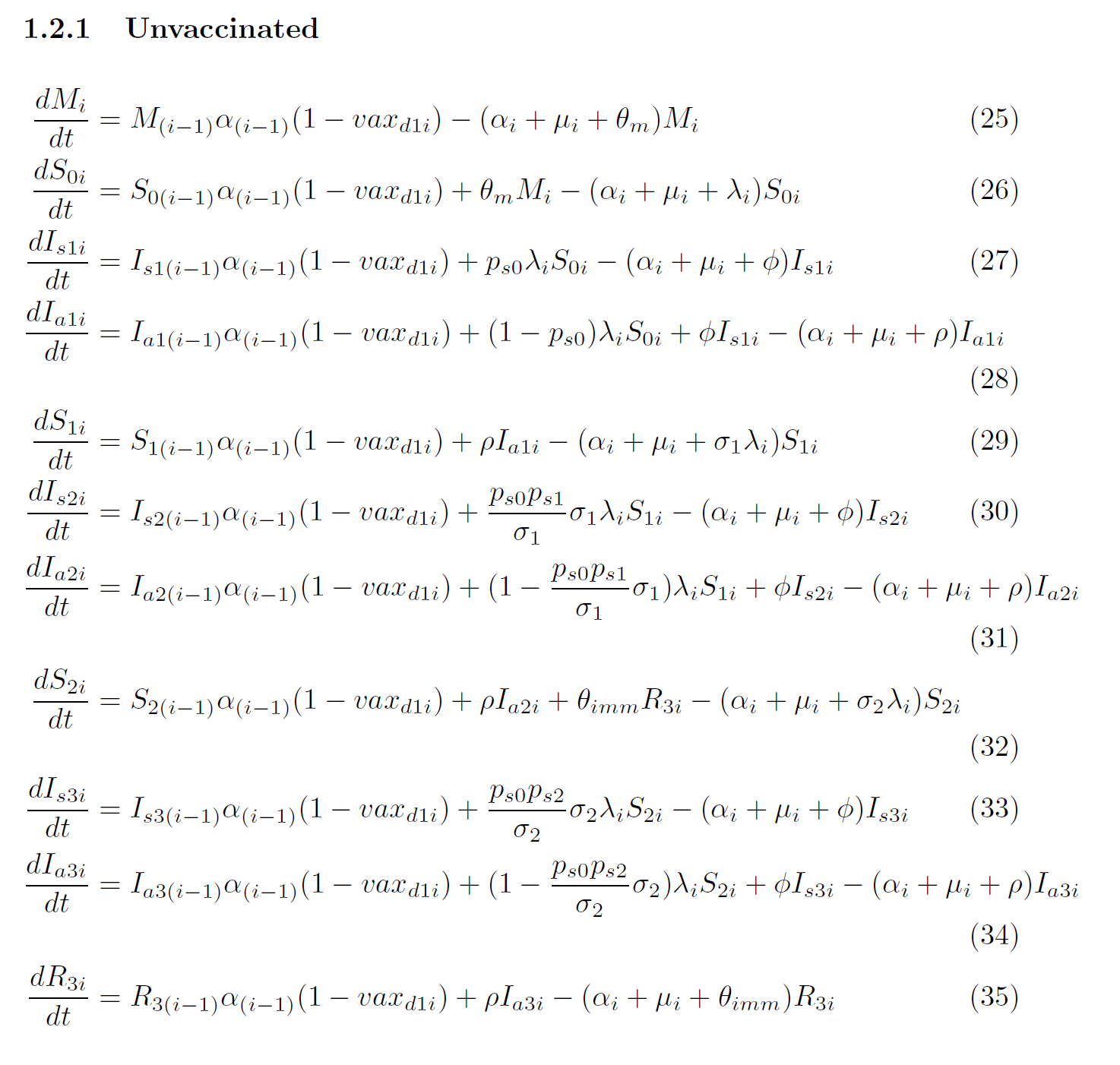

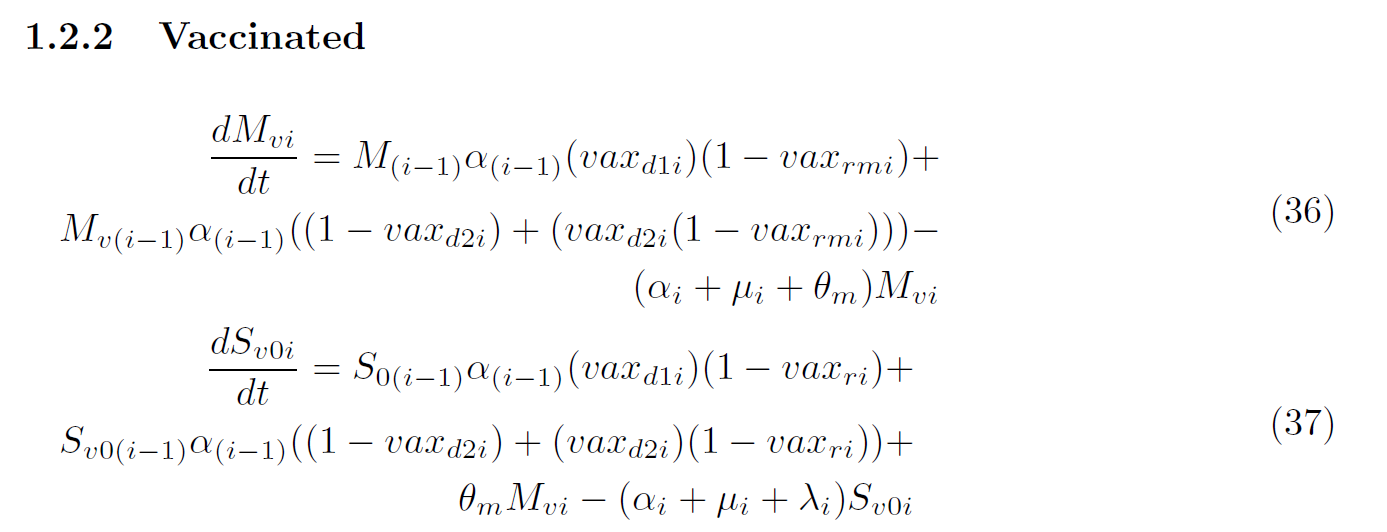

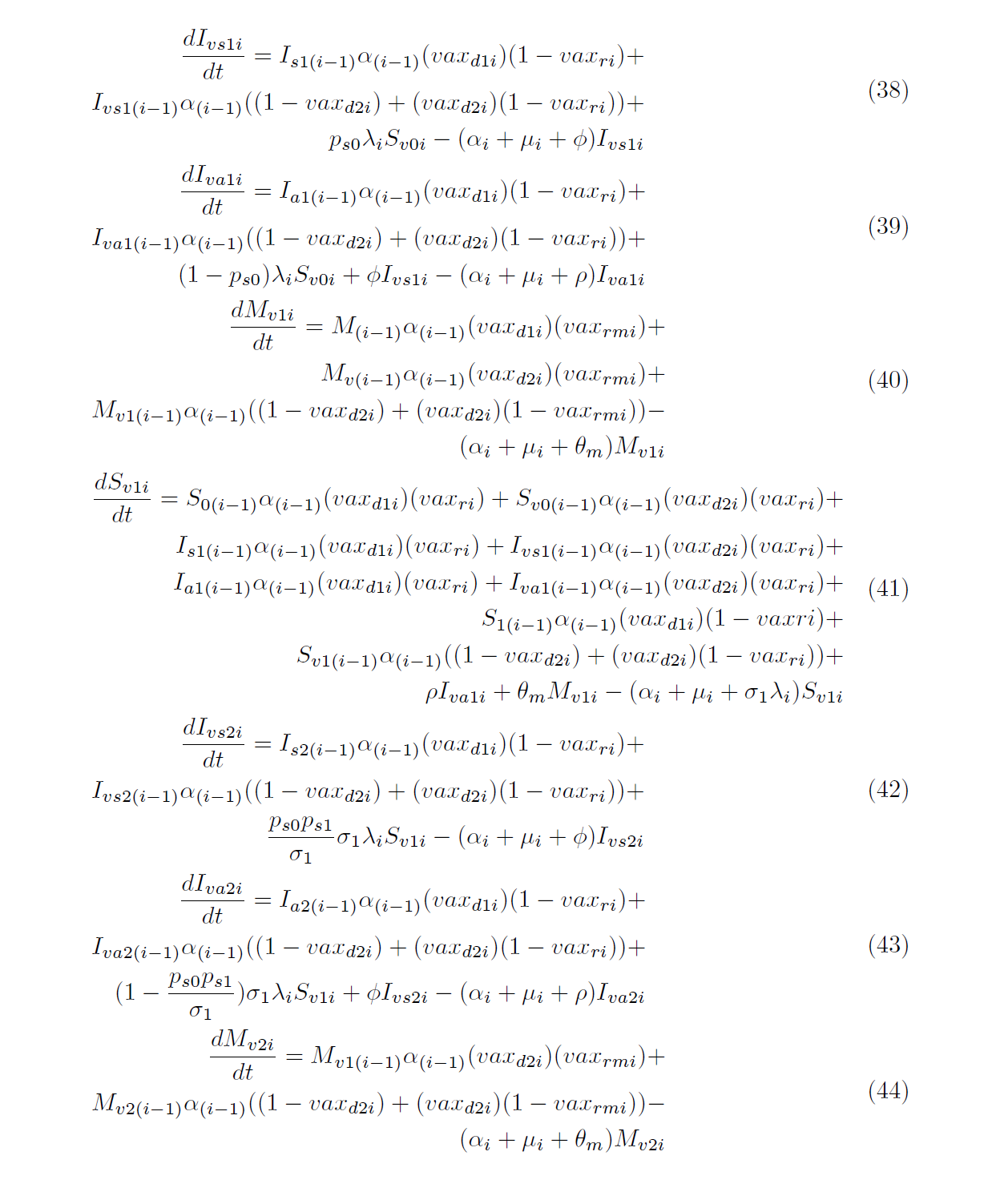

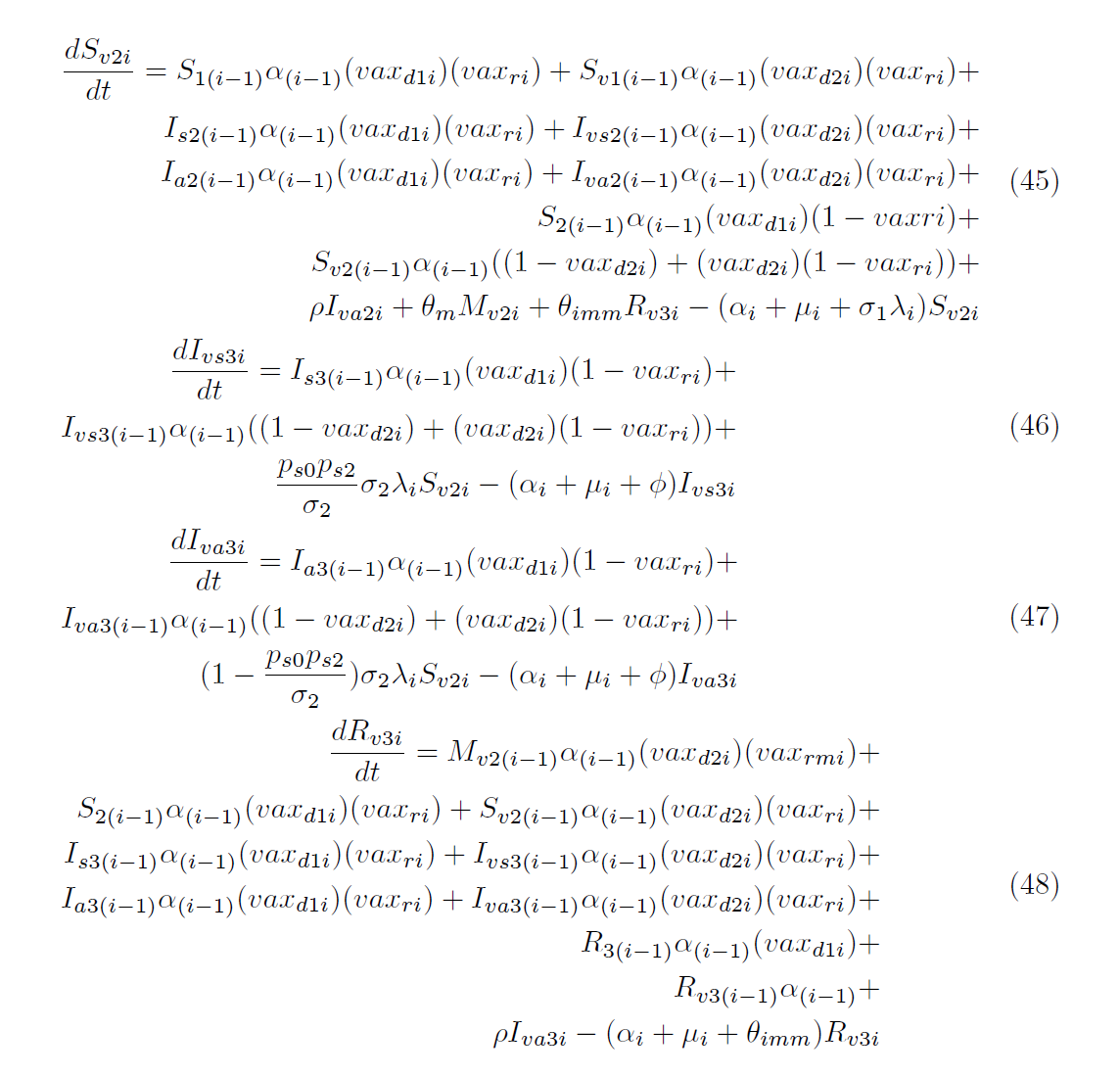

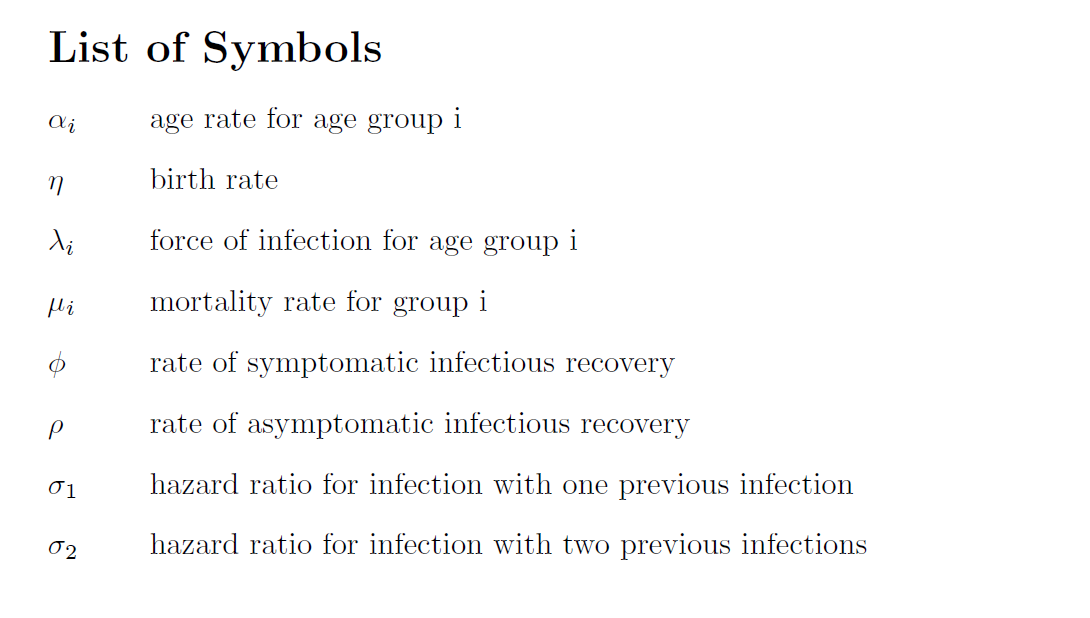

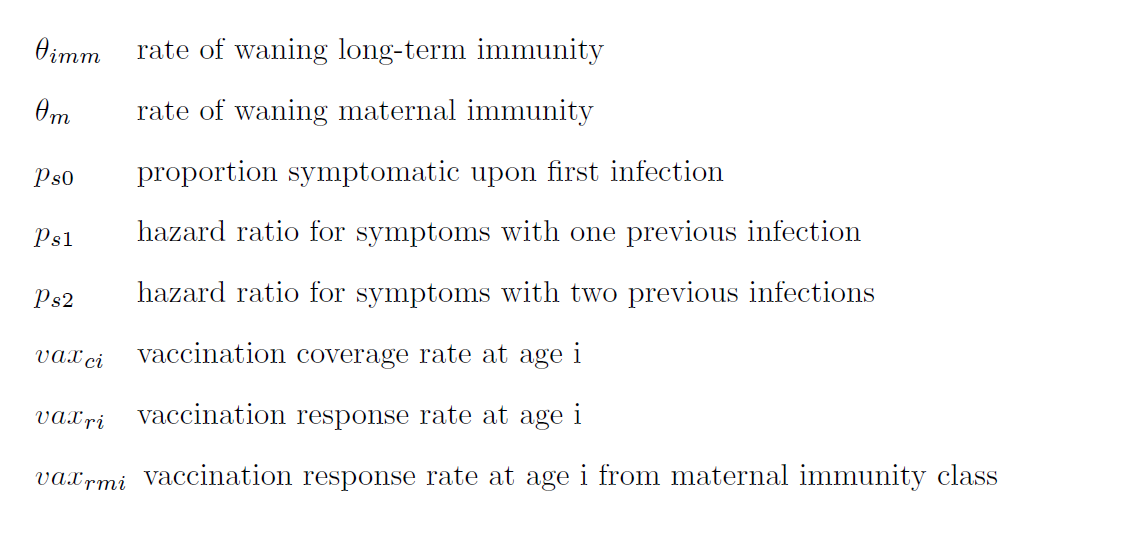

3. **Population mixing and transmission rates**

We assumed population mixing and contact rates varied between age groups and was homogenous within age groups. Contact rates were calculated from the raw POLYMOD data from 8 European countries (Belgium, Germany, Finland, Great Britain, Italy, Luxembourg, The Netherlands, and Poland) for five age groups: infants (<1 years old), young children (1 - <5 years old), older children (5 - <18 years old), adults (18 - <65 years old), older adults (65+ years old), as described in Steele et al.^1,2^ The total daily contacts each age group makes are:

<1 year olds (c_1_) = 5.43

1 - <5 year olds (c_2_) = 8.57

5 - <18 year olds (c_3_) = 15.32

18 - <65 year olds (c_4_) = 14.27

65+ year olds (c_5_) = 8.53

These daily contact rates are then used to calculate the proportion of daily contacts an age group makes with each other age group, per Steele et al^1^.

The force of infection incorporates age-specific contact rates, seasonality, and norovirus transmissibility. Seasonality is incorporated as a sinusoidal function, σ(t), as follows:

$$\sigma\left( t \right)=1+ \beta_{1} \times\cos\left( 2\pi t+\omega\right)$$

where β_1_ represents the amplitude of seasonal forcing and ω represents the seasonal offset parameter.

The force of infection is calculated as follows:

$$\lambda_{i}\left( t \right)=b_{i}\beta_{0i}\sigma\left( t \right)\sum_{j=1}^{5} c_{ij}\frac{\left( I_{j}+\varepsilon A_{j} \right)}{N_{j}}$$

where *b_i_* is the total number of contacts made by and individual in age group *i* per day*,* $\beta_{0i}$ represents the age-specific transmission probability of norovirus (transmission probability age groups differ from age groups and include <5 years old, 5 – 64 years old, 65+ years old), *c_ij_* is the proportion of contacts individuals in age group *i* make with age group *j*, ε represents the relative infectiousness of asymptomatic individuals compared to symptomatic individuals, N_j_ represents population size of age group j, and I_j_ and A_j_­ represent the number of individuals of age group j infected symptomatically and asymptomatically, respectively, such that $\frac{\left( I_{j}+\varepsilon A_{j} \right)}{N_{j}}$ represents the infectious fraction of age group j.

4. **Model fitting**

In this model, we estimated best-fit values for the age-specific transmission parameters (β­_0k_), seasonality coefficients (β­_1_, ω), rate of waning of maternal immunity (μ), rate of waning of long-term immunity (θ), and proportion symptomatic upon initial infection (p_s0_) by fitting the age group-specific model-predicted weekly reported cases, *m_it_,* to the observed number of weekly reported cases by age group, ­*y_it_*.*^.^*

In this study, we were most interested in most closely representing early life infection and disease dynamics in order to better capture the impact of and compare differences in pediatric norovirus vaccination strategies. When fitting the model, we aimed to capture these dynamics by fitting to smaller age groups among younger children with larger age groups among older children and adults: <6m, 6m - <1 yr, 1yr - <2 yr, 2yr - <3 yr, 3yr - <4yr, 4yr - <5yr, 5 - <18yr, 18 - <65yr, >65yr.

Observed number of reported and lab-confirmed German norovirus cases for single year age groups were acquired from the Robert Koch Incident SurvStat database^3^, which includes weekly case counts. Case counts were summed into larger age groups as necessary and scaled to the US population, using population data from the UN World Population Prospects (Germany) and CDC Wonder database (US)^4,5^, as follows:

$$y_{it}=({reported cases}_{it}*\frac{{pop}_{US,i}}{{pop}_{Germany,i}})$$

where ­*y_it_* is the number of total cases in the US in age group *i* at time *t*, ${reported cases}_{i}$ is the number of reported cases in Germany for age group *i* at time *t*, *pop­_US,i_* and *pop_Germany,i_* are the population size for age group *i* in the US and Germany, respectively (assumed to be constant over time).

In the case of the <1 population, case counts were divided by <6m and 6m - <1yr based on the estimated proportion of pediatric norovirus cases occurring prior to 6m of age from a global meta-analysis^6^.

We calculated the model-predicted daily reported cases for each age group, which was summed by week, as follows:

$$m_{it}=r\sum_{t} ( p_{s0}\lambda_{i,t}S_{0i,t}+ \frac{p_{s0}p_{s1}}{\sigma_{1}}{\sigma_{1} \lambda}_{i,t}S_{1i,t}+ \frac{p_{s0}p_{s2}}{\sigma_{2}}\sigma_{2}\lambda_{i,t}S_{2i,t} )$$

where $r$ is the norovirus case reporting rate. The reporting rate was calculated as follows:

$$r= \frac{1}{expected cases} \sum_{i} \sum_{t} y_{it}$$

where cases were summed annually (*t*) and by age group (*i*) and the expected cases are the number of annual norovirus cases estimated for the US across all age groups (20 million)^7^.

For each individual age group (*i*), the negative-log likelihood (NLL) was calculated by assuming the reported weekly cases were Poisson distributed with the mean equal to the model-predicted reported weekly cases. The age-specific NLL were combined into an overall NLL, which was minimized to obtain the best-fit parameters and model fit:

$$LL=\sum_{i} \sum_{t} \left( y_{i,t}\log\left( m_{i,t} \right)-m_{i,t} \right)$$

**Supplemental Figures and Tables**

**Figure 1: Observed (red) and best fit model-predicted (blue) reported weekly norovirus cases by age group to assess model fit.** Peak in observed cases (particularly notable in the young children, older children, and adults) is from the 2016 winter season and may be associated with the emergence of a new recombinant norovirus strain, GII.P16-GII.2^8^.

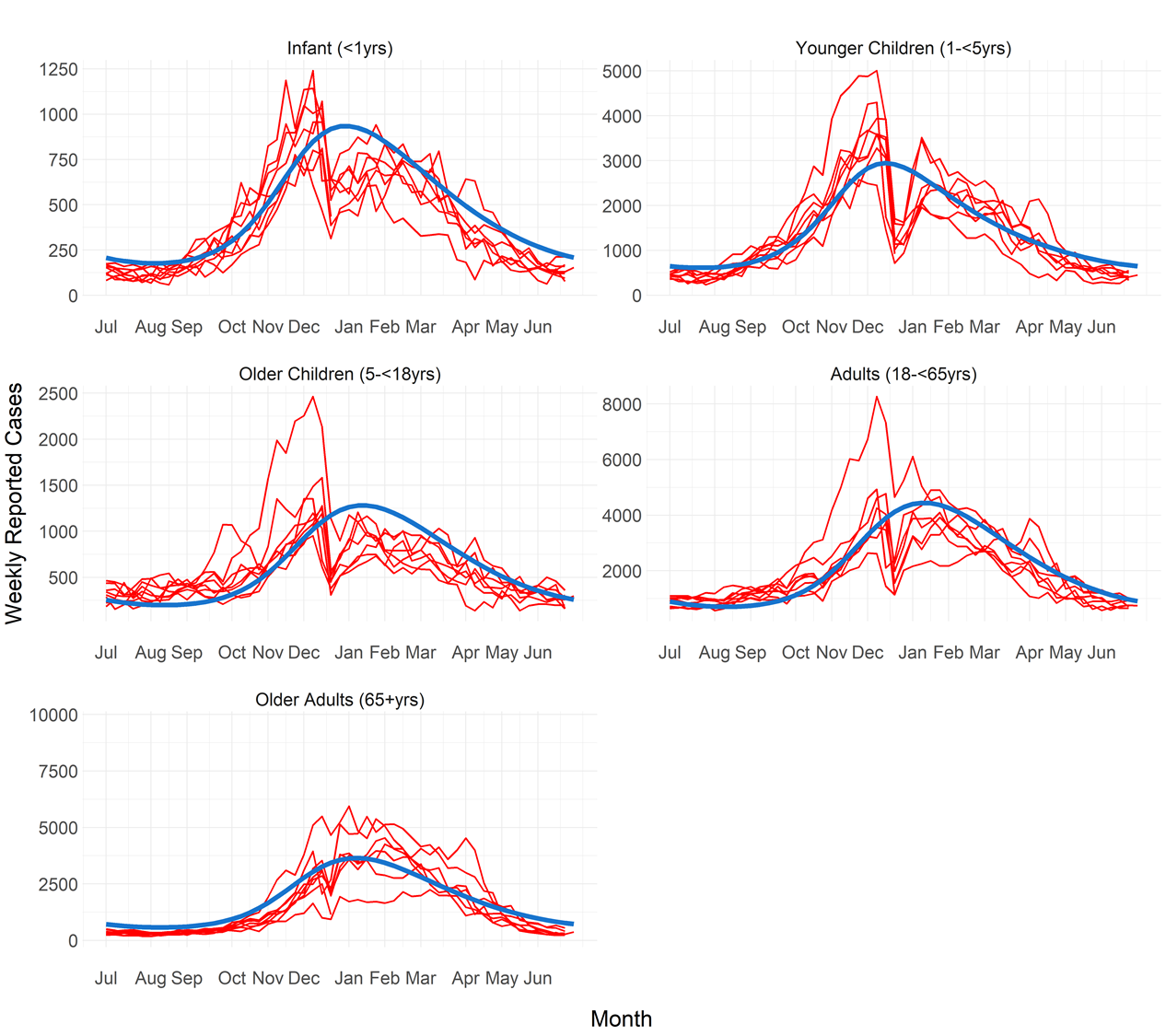

**Figure 2: Norovirus AGE cases averted for five years following the introduction of vaccination given 70% vaccination coverage and with maternal antibody interference** as (a) annual cases averted per 100,000 and (b) weekly cases averted per 100,000, for young children (<5 years of age) and total population across vaccination schedules.

**A**

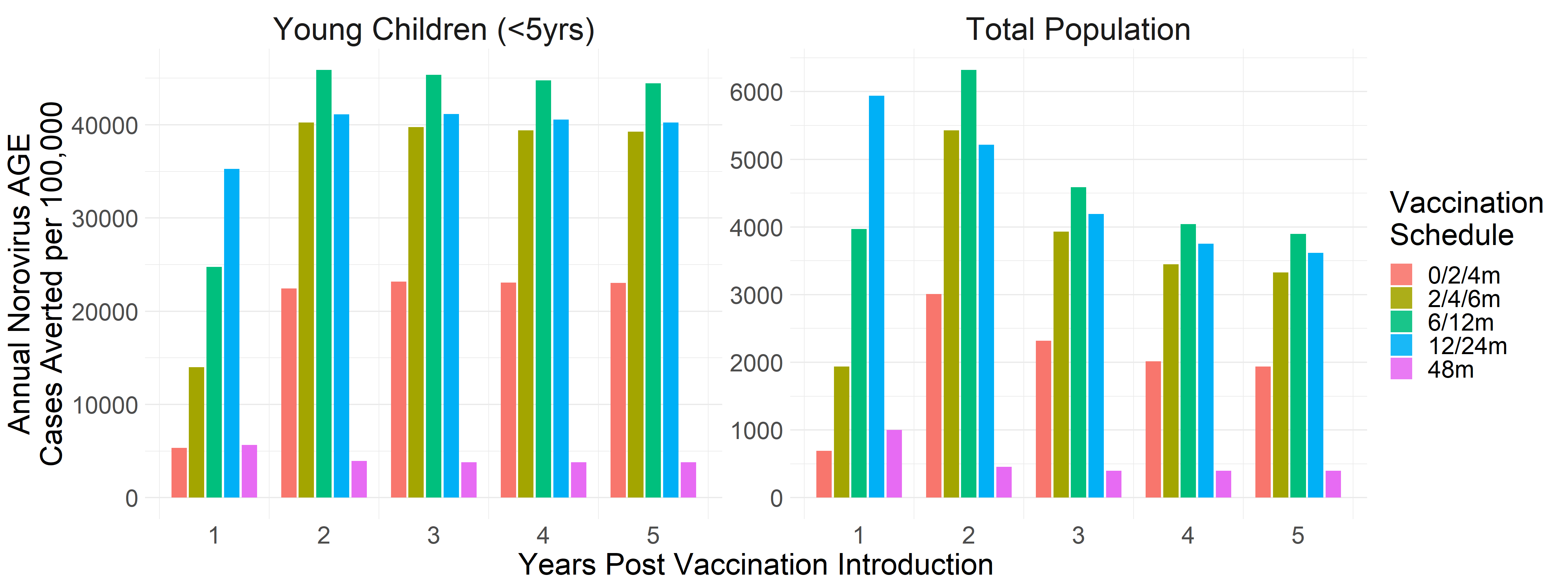

**B**

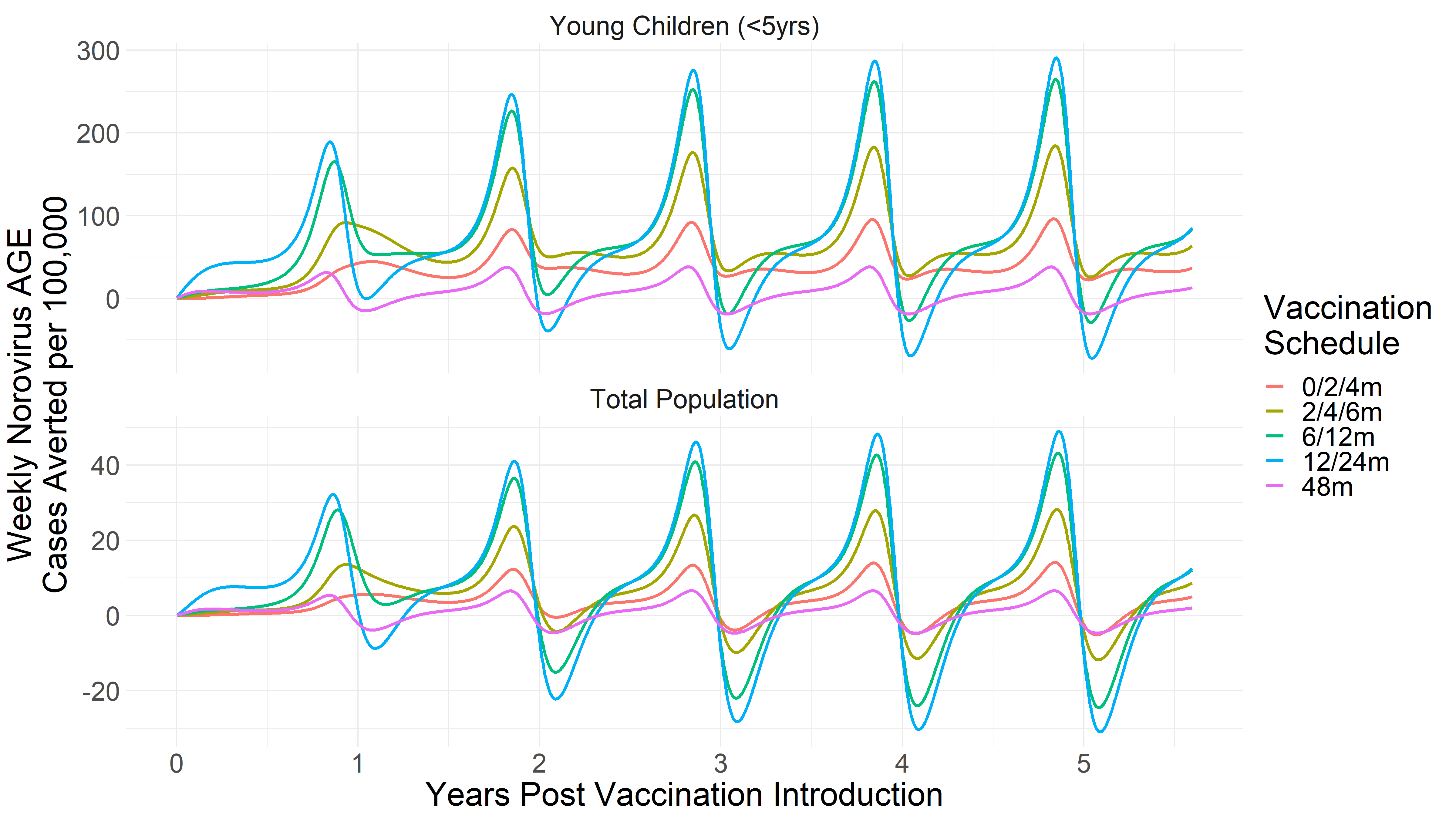

**Figure 3: Norovirus AGE cases averted for five years following the introduction of vaccination given 90% vaccination coverage and no maternal antibody interference** as (a) annual cases averted per 100,000 and (b) weekly cases averted per 100,000, for young children (<5 years of age) and total population across vaccination schedules.

**A**

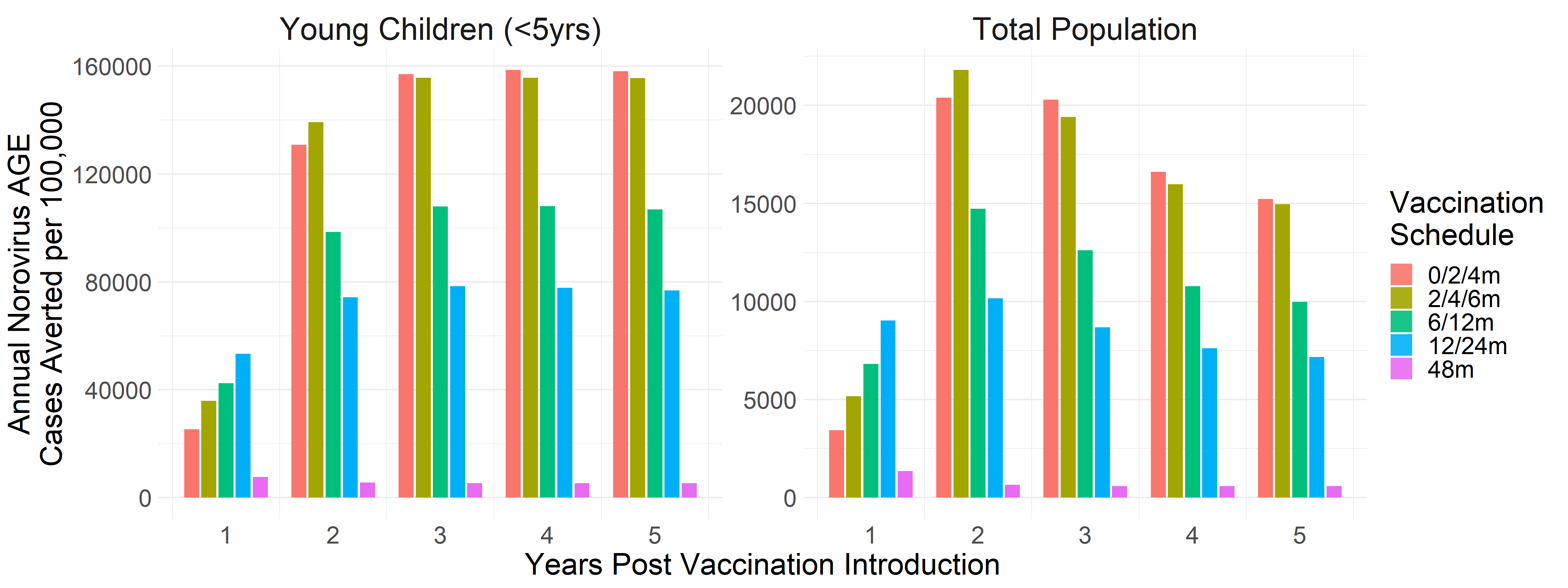

**B**

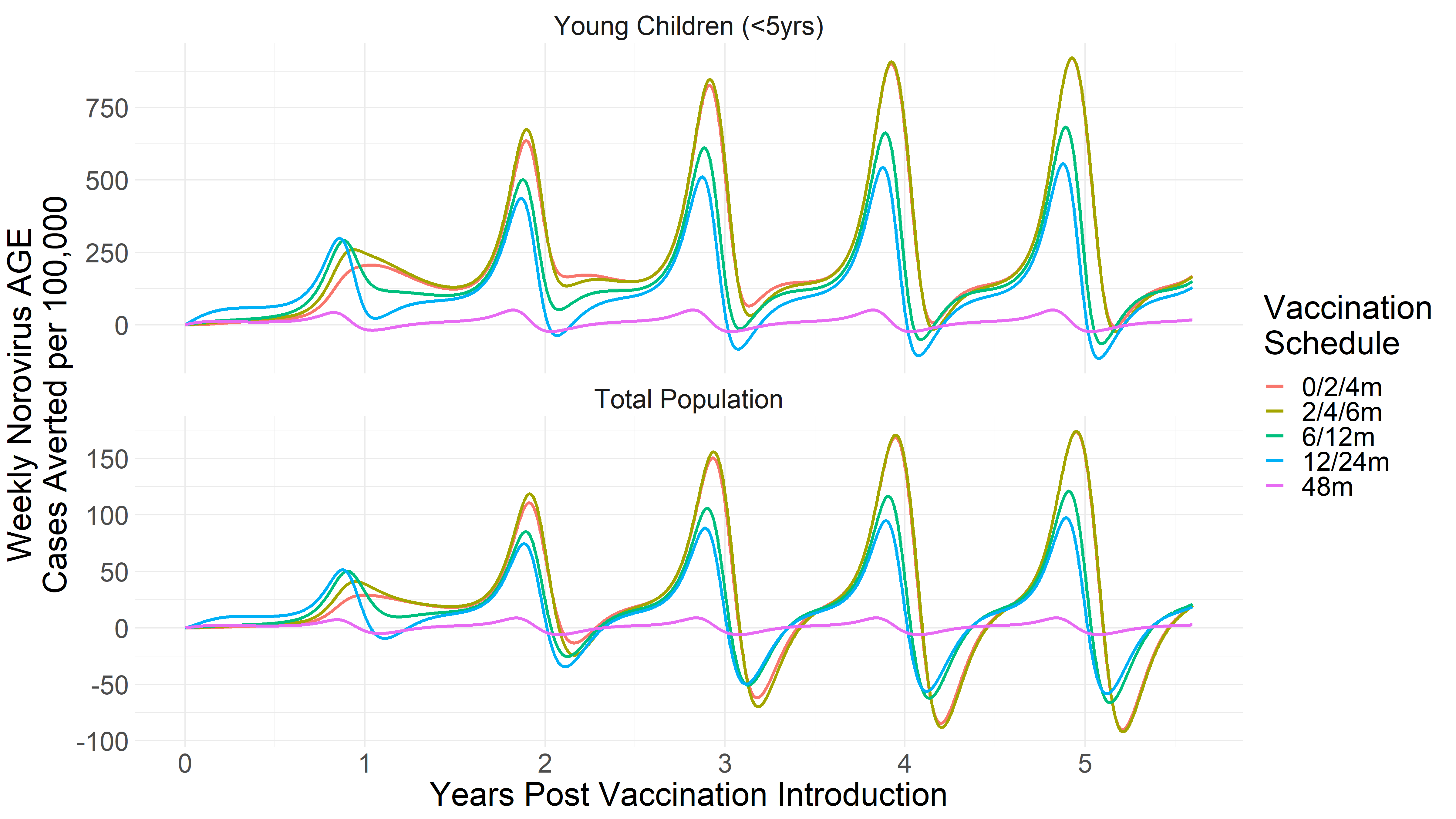

**Figure 4: Norovirus AGE cases averted for five years following the introduction of vaccination given 90% vaccination coverage and with maternal antibody interference** as (a) annual cases averted per 100,000 and (b) weekly cases averted per 100,000, for young children (<5 years of age) and total population across vaccination schedules.

**A**

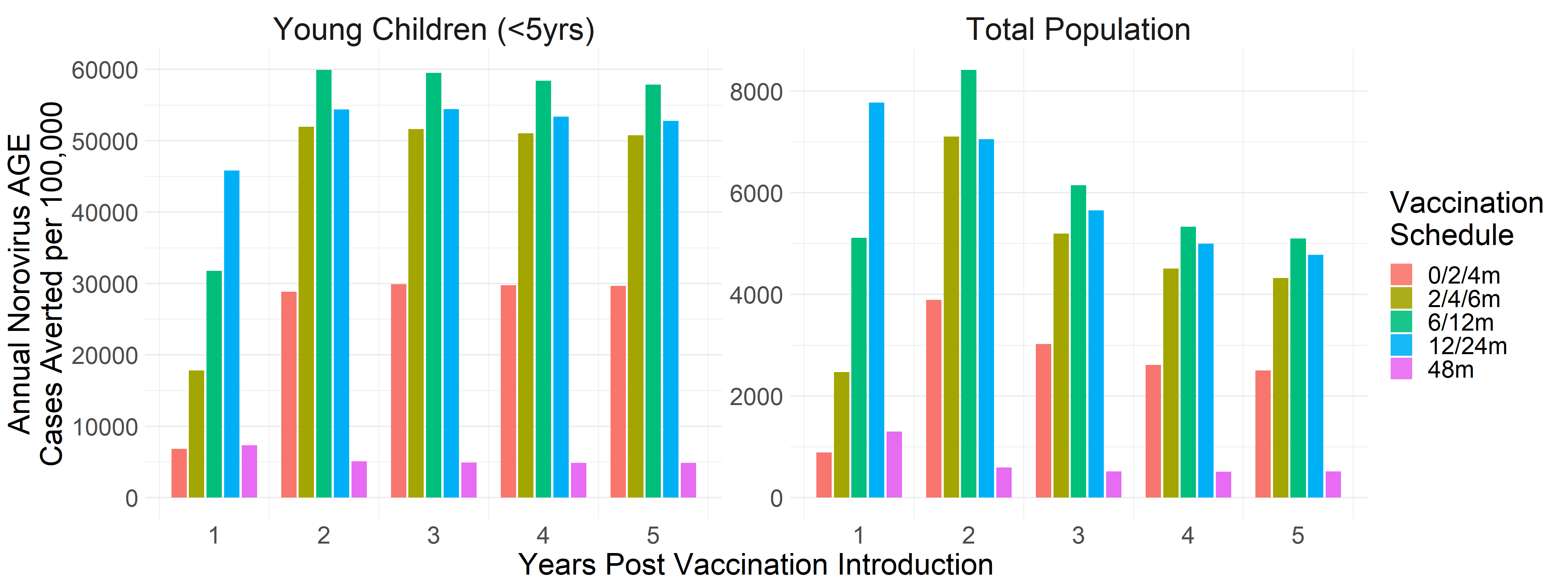

**B**

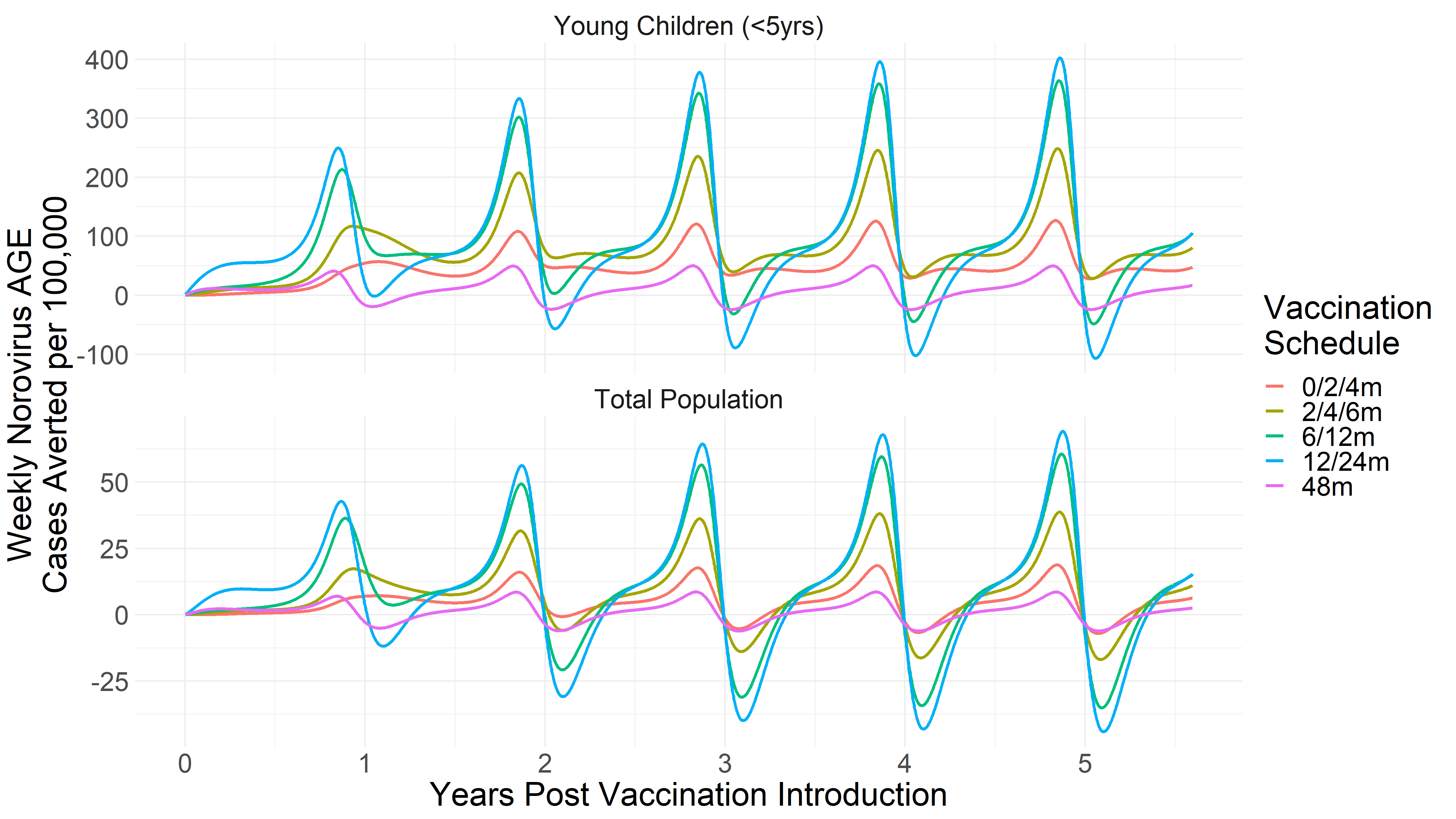

**Figure 5: Annual direct and indirect effects of pediatric norovirus vaccination as percent norovirus AGE cases averted, given 90% vaccination coverage** (a) with no maternal antibody interference with vaccine efficacy and (b) with complete maternal antibody interference with vaccine efficacy: Different vaccine schedules are indicated by bar colors. Direct vs indirect effects represented by dark vs light opacity. Each plot represents the annual percent cases averted for a single year after five years of vaccination. Percent cases averted are presented for individual age groups and the total population.

**A**

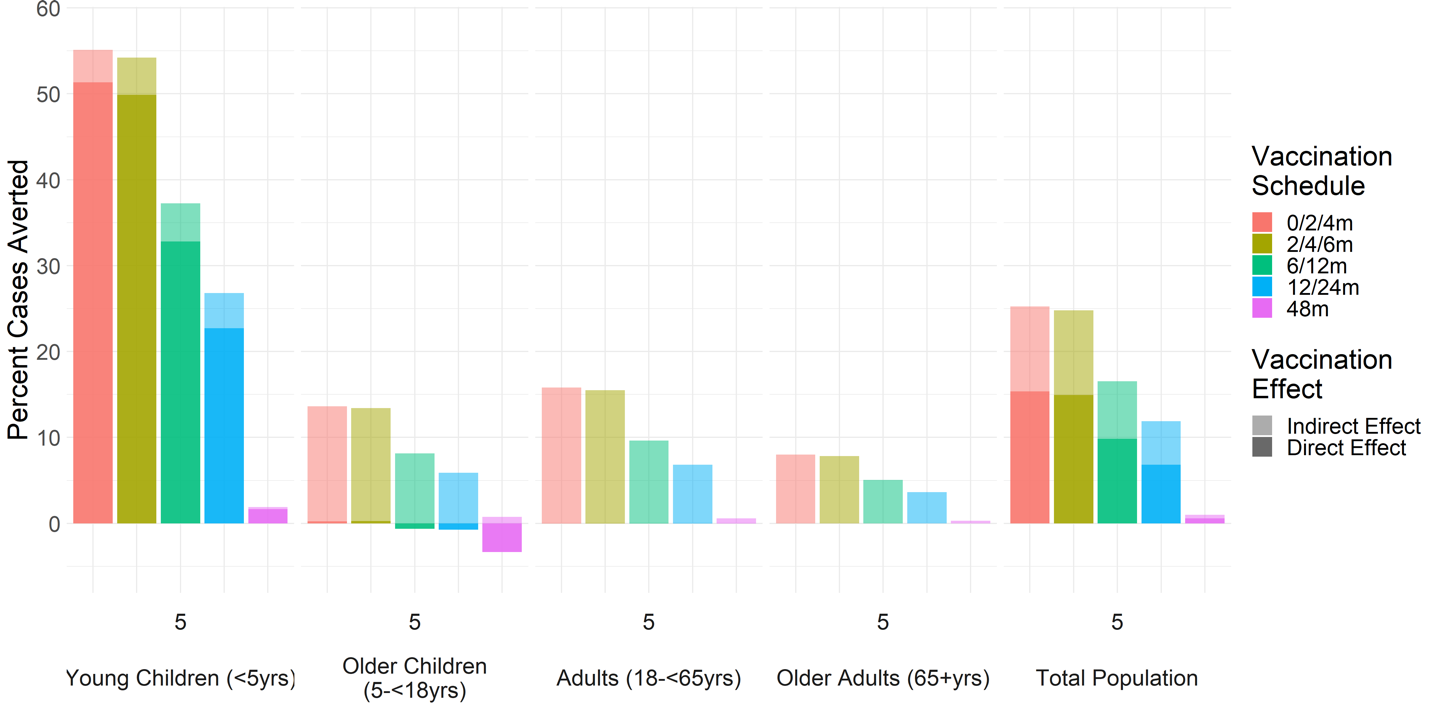

**B**

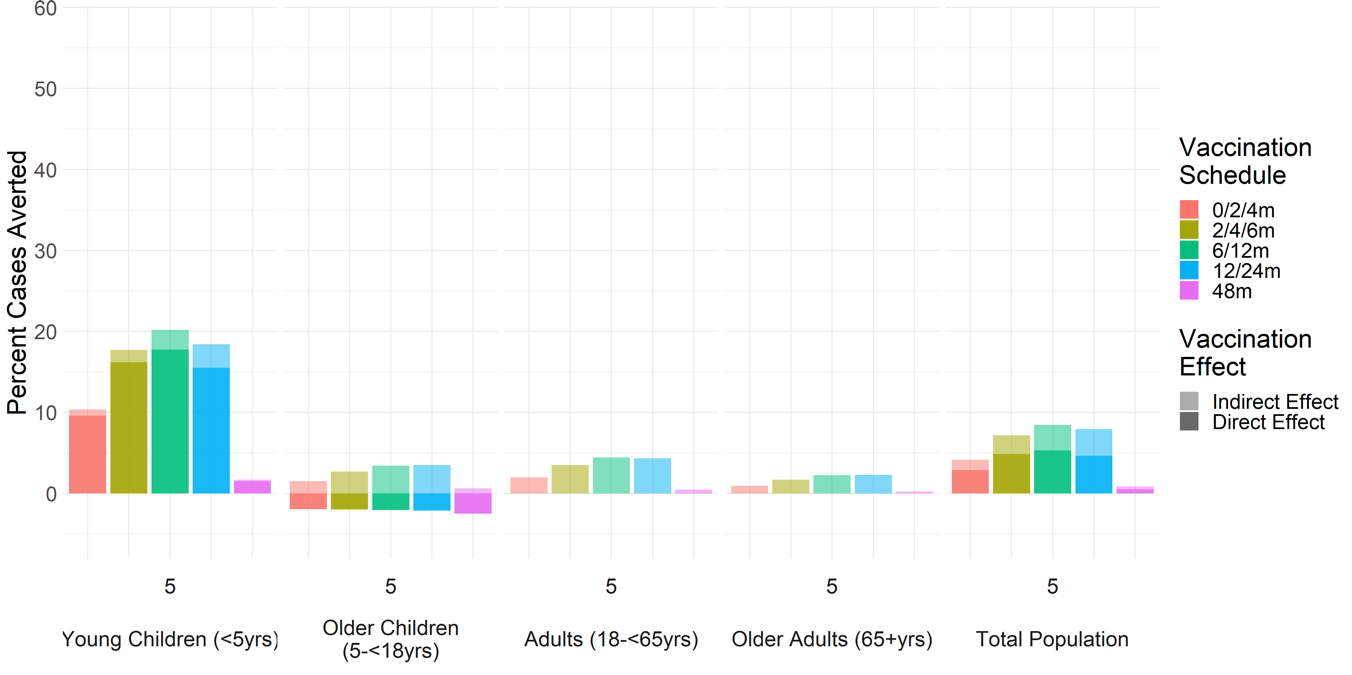

**Figure 6: Sensitivity Analysis: Boxplots of annual percent norovirus AGE cases averted with vaccination a) without maternal antibody interference with vaccination and b) with maternal antibody interference with vaccination.** Results based on 1000 parameter draws generated using Latin Hypercube Sampling of parameter distributions provided in Table 1. Different vaccine schedules are indicated by box colors. Percent cases averted are presented for individual age groups and the total population and for 70% vaccination coverage.

**A**

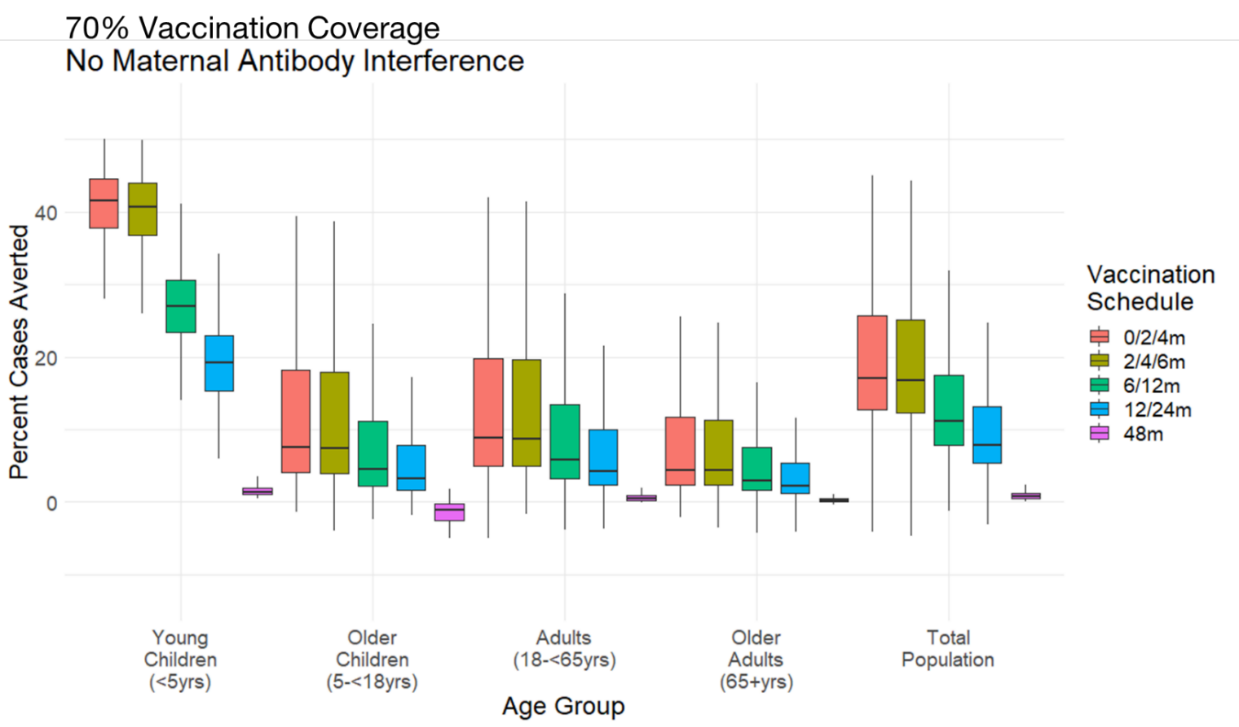

**B**

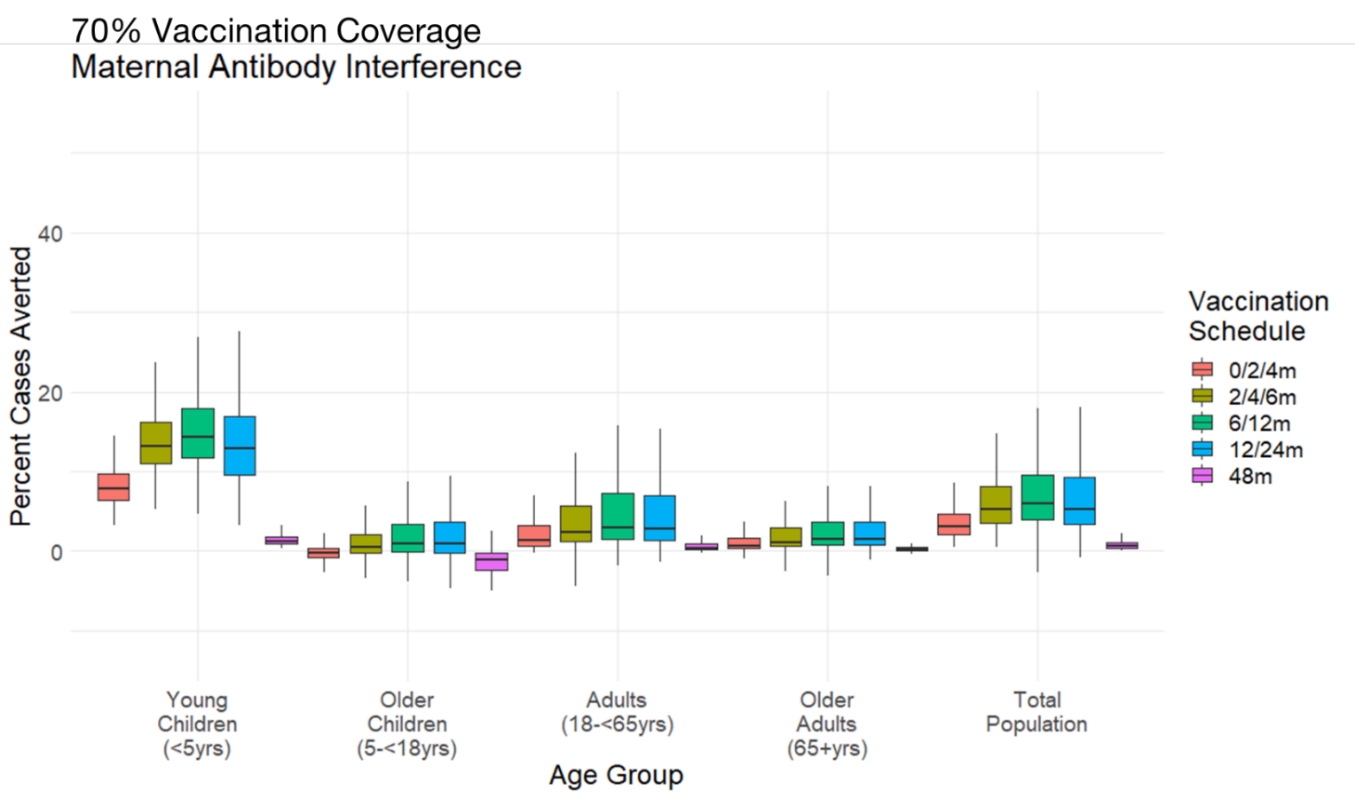

C

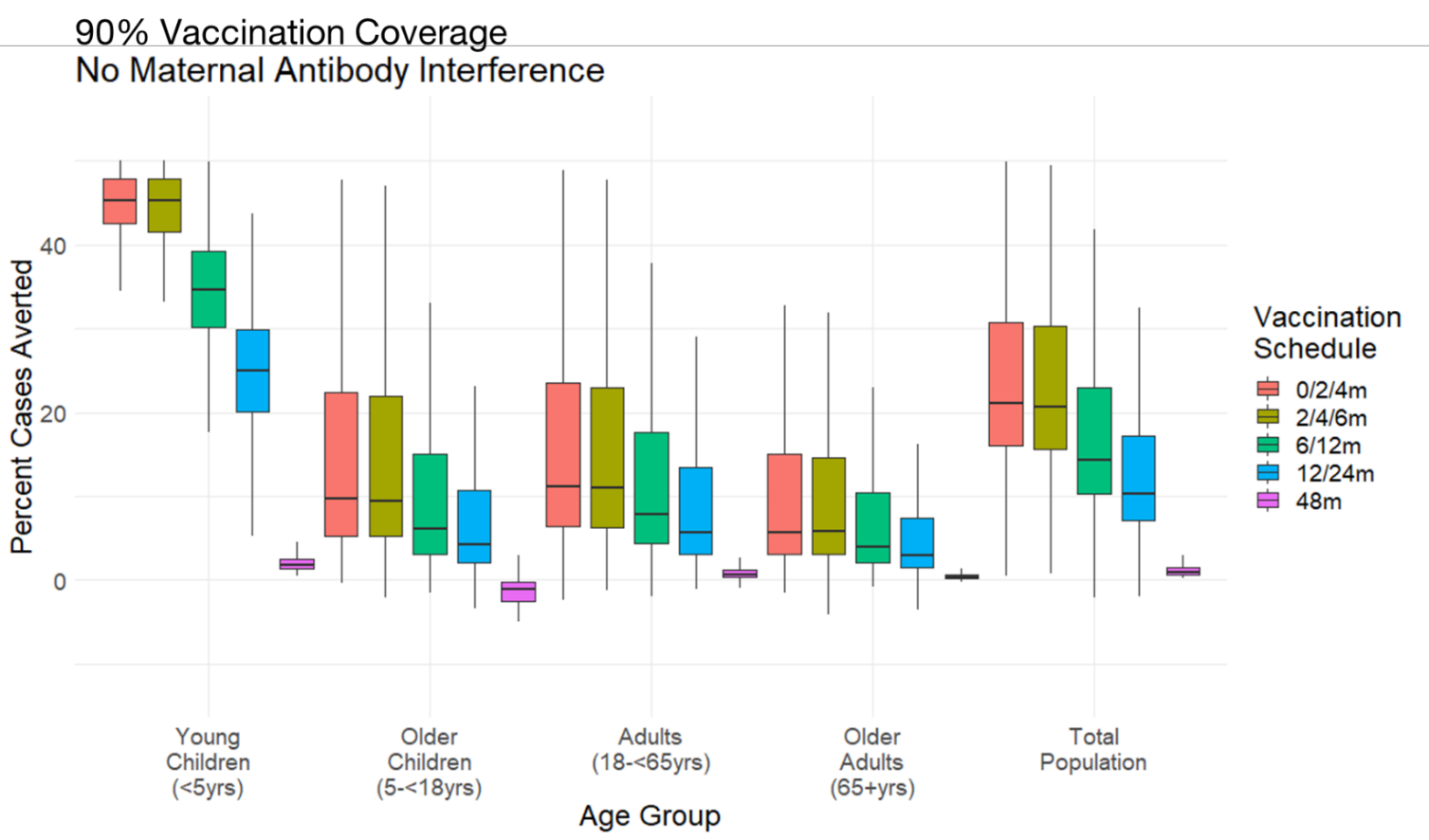

**D**

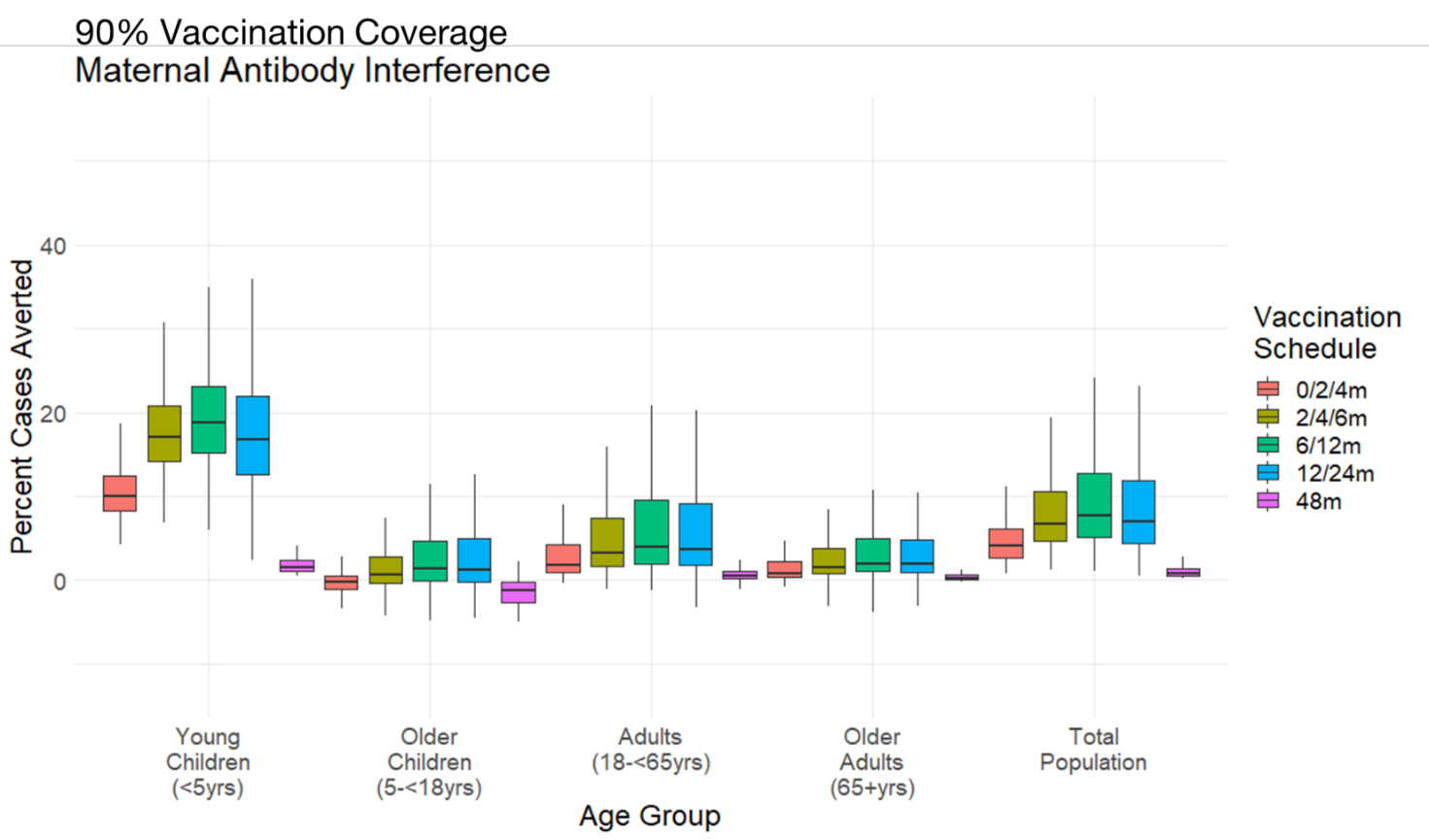

**Figure 7: Sensitivity Analysis: Partial rank correlation coefficient (PRCC) for model parameters associated with percent cases averted a) in <5 year olds and b) in the total population.** Percent cases averted distributions were based on 1000 parameter draws generated using Latin Hypercube Sampling of parameter distributions provided in Table 1 and 70% vaccination coverage. Bar colors represent PRCC associated with and without maternal antibody interference. PRCC calculated using 100 bootstrap replicates.

**A**

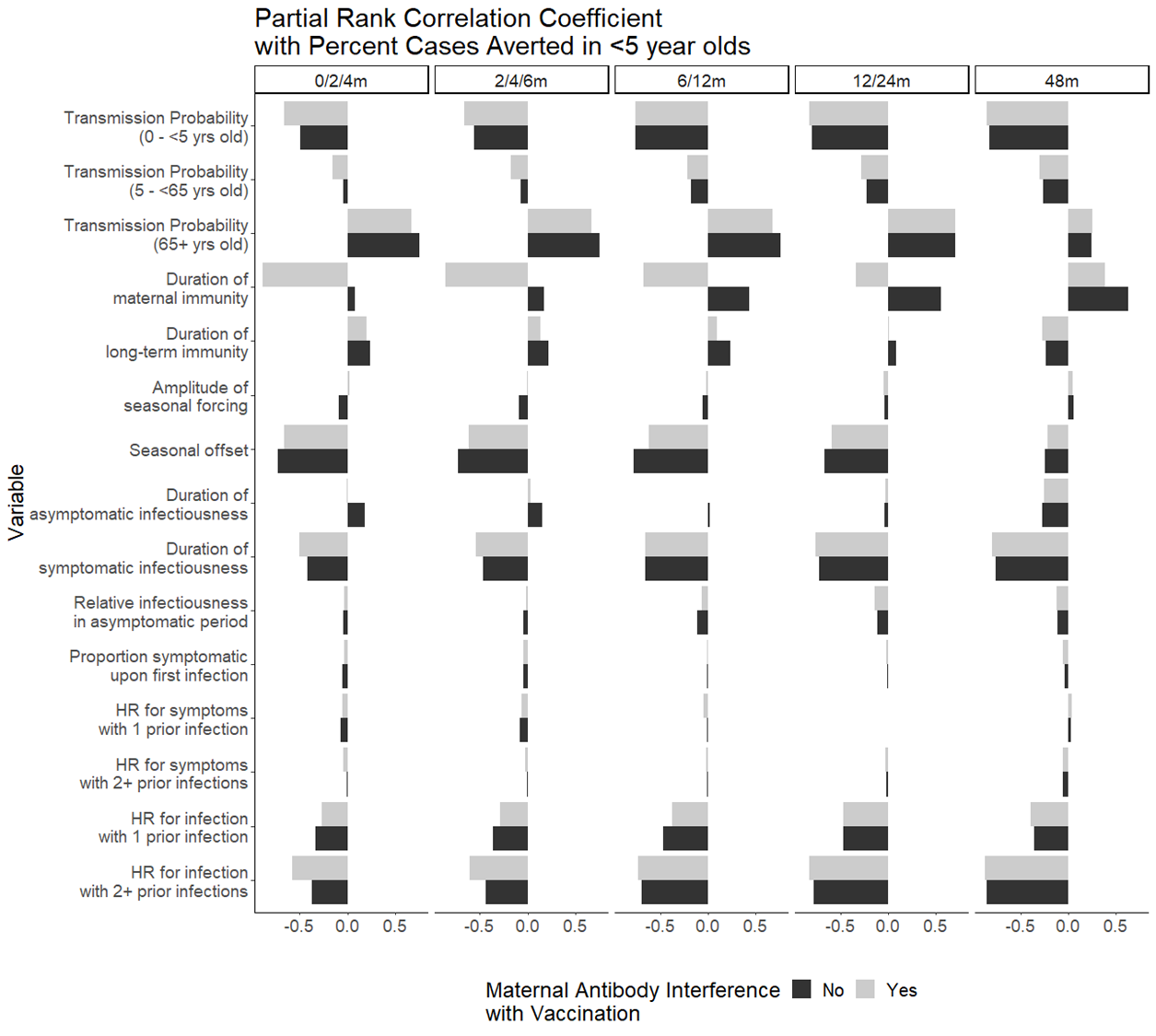

**B**

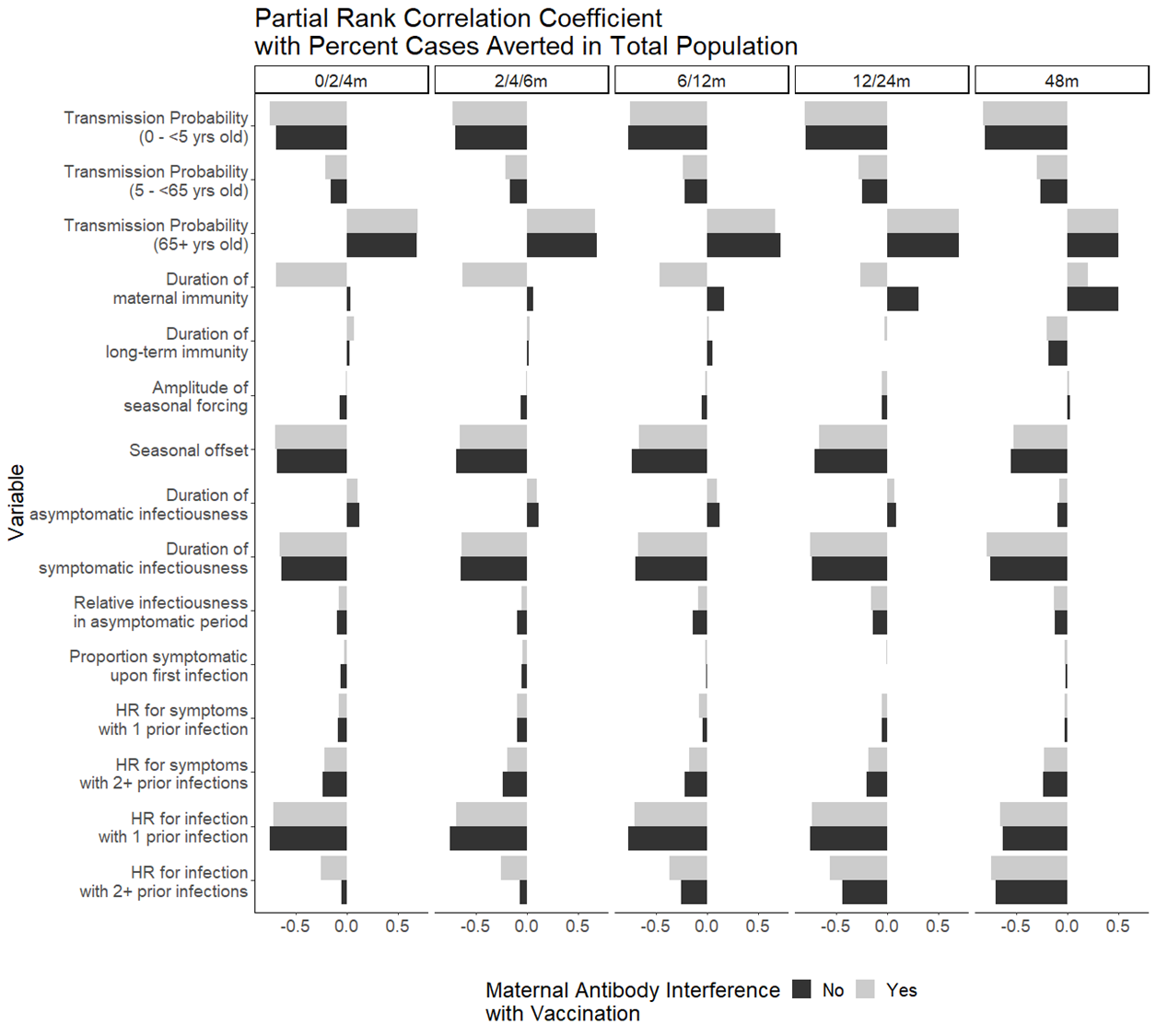

**Table 1: Total clinical outcomes averted annually by vaccination scenario (95% uncertainty interval based on LHS sensitivity analysis) by age group.** Results associated with maternal antibody interference with vaccine efficacy and 70% vaccination coverage.

| *Age Group* | *Clinical Outcome* | *Outcomes Averted by Vaccination Scenario  (95% UI)* | | | | |
| --- | --- | --- | --- | --- | --- | --- |
|  |  | **0/2/4m** | **2/4/6m** | **6/12m** | **12/24m** | **48m** |
| *Young Children (<5yrs)* | **AGE Cases** | 470000  (250000 - 1100000) | 800000  (430000 - 1840000) | 910000  (490000 - 1810000) | 830000  (420000 - 1680000) | 80000  (40000 - 230000) |
|  | **Outpatient Visits** | 79000  (35000 - 204000) | 135000  (60000 - 348000) | 153000  (68000 - 360000) | 139000  (55000 - 322000) | 13000  (5000 - 42000) |
|  | **Hospitalizations** | 2000  (1100 - 4800) | 3400  (1800 - 8000) | 3900  (2100 - 8000) | 3500  (1800 - 7300) | 300  (200 - 1000) |
|  | **Deaths** | 3  (2 - 7) | 5  (3 - 12) | 6  (3 - 12) | 5  (3 - 11) | 0  (0 - 1) |
|  | **Percent Outcomes Averted*** | 8%  (4 - 15%) | 14%  (7 - 25%) | 16%  (7 - 29%) | 14%  (5 - 31%) | 1%  (1 - 6%) |
| *Total  Population* | **AGE Cases** | 630000  (340000 - 1510000) | 1090000  (590000 - 2640000) | 1270000  (650000 - 2920000) | 1180000  (590000 - 2730000) | 130000  (60000 - 480000) |
|  | **Outpatient Visits** | 88000  (39000 - 233000) | 154000  (68000 - 400000) | 179000  (77000 - 441000) | 164000  (64000 - 406000) | 11000  (-15000 - 52000) |
|  | **Hospitalizations** | 2900  (1600 - 8200) | 5000  (2600 - 14300) | 6000  (2900 - 16600) | 5600  (2600 - 16600) | 500  (100 - 2600) |
|  | **Deaths** | 20  (8 - 102) | 35  (14 - 181) | 46  (16 - 217) | 46  (15 - 228) | 5  (1 - 41) |
|  | **Percent AGE Cases Averted** | 3%  (1 - 12%) | 6%  (2 - 21%) | 6%  (2 - 24%) | 6%  (1 - 26%) | 1%  (0 - 5%) |
|  | **Percent Outpatient Visitis Averted** | 4%  (1 - 12%) | 7%  (2 - 22%) | 8%  (2 - 24%) | 7%  (2 - 27%) | 0%  (0 - 4%) |
|  | **Percent Hospitalizations Averted** | 2%  (1 - 10%) | 4%  (1 - 18%) | 4%  (1 - 21%) | 4%  (1 - 23%) | 0%  (0 - 4%) |
|  | **Percent Deaths Averted** | 1%  (0 - 8%) | 1%  (0 - 14%) | 2%  (0 - 18%) | 2%  (0 - 20%) | 0%  (0 - 3%) |
|  | **Doses to avert a case**** | 13.3  (6 - 25.5) | 7.5  (3.4 - 14.2) | 4.1  (1.9 - 7.8) | 4.1  (1.9 - 7.9) | 18.9  (5.5 - 40.8) |
|  | **Vaccinees to avert a case**** | 4.5  (2.1 - 8.8) | 2.6  (1.2 - 4.9) | 2.1  (1 - 4.1) | 2.2  (1 - 4.4) | 18.9  (5.5 - 40.8) |
|  | **Cases averted per dose**** | 0.1  (0 - 0.2) | 0.1  (0.1 - 0.3) | 0.2  (0.1 - 0.5) | 0.2  (0.1 - 0.5) | 0.1  (0 - 0.2) |
|  | **Cases averted per vaccinee**** | 0.2  (0.1 - 0.5) | 0.4  (0.2 - 0.8) | 0.5  (0.2 - 1) | 0.5  (0.2 - 1) | 0.1  (0 - 0.2) |
| *Notes:* | *As clinical outcomes are scaled from AGE cases, the percent outcomes averted are the same across outcomes within an age group. For the total population, individual percent outcomes averted are presented for each clinical outcome. | | | | | |

**Table 2: Vaccine efficiency by vaccination scenario (95% uncertainty interval based on LHS sensitivity analysis) by age group with maternal antibody interference with vaccine efficacy and 70% vaccination coverage.** Vaccine efficiency measures are presented for the cumulative doses or vaccinated individuals and cumulative cases averted over five years.

| *Efficiency Measure* | *Efficiency Value by Vaccination Scenario  (95% UI)* | | | | |
| --- | --- | --- | --- | --- | --- |
|  | **0/2/4m** | **2/4/6m** | **6/12m** | **12/24m** | **48m** |
| Doses to avert a case | 13.3  (6 - 25.5) | 7.5  (3.4 - 14.2) | 4.1  (1.9 - 7.8) | 4.1  (1.9 - 7.9) | 18.9  (5.5 - 40.8) |
| Vaccinees to avert a case | 4.5  (2.1 - 8.8) | 2.6  (1.2 - 4.9) | 2.1  (1 - 4.1) | 2.2  (1 - 4.4) | 18.9  (5.5 - 40.8) |
| Cases averted per dose | 0.1  (0 - 0.2) | 0.1  (0.1 - 0.3) | 0.2  (0.1 - 0.5) | 0.2  (0.1 - 0.5) | 0.1  (0 - 0.2) |
| Cases averted per vaccinee | 0.2  (0.1 - 0.5) | 0.4  (0.2 - 0.8) | 0.5  (0.2 - 1) | 0.5  (0.2 - 1) | 0.1  (0 - 0.2) |

**Table 3: Total clinical outcomes averted annually by vaccination scenario (95% uncertainty interval based on LHS sensitivity analysis) by age group with no maternal antibody interference with vaccine efficacy and 90% vaccination coverage.**

| *Age Group* | *Clinical Outcome* | *Outcomes Averted by Vaccination Scenario  (95% SI)* | | | | |
| --- | --- | --- | --- | --- | --- | --- |
|  |  | **0/2/4m** | **2/4/6m** | **6/12m** | **12/24m** | **48m** |
| *Young Children (<5yrs)* | **AGE Cases** | 3240000  (1910000 - 5860000) | 3190000  (1870000 - 5740000) | 2190000  (1260000 - 3710000) | 1580000  (850000 - 2870000) | 110000  (50000 - 310000) |
|  | **Outpatient Visits** | 545000  (245000 - 1220000) | 536000  (242000 - 1192000) | 368000  (168000 - 772000) | 265000  (115000 - 570000) | 19000  (7000 - 57000) |
|  | **Hospitalizations** | 13900  (8200 - 25400) | 13600  (7900 - 24900) | 9400  (5400 - 16100) | 6700  (3600 - 12500) | 500  (200 - 1400) |
|  | **Deaths** | 20  (12 - 37) | 20  (12 - 36) | 14  (8 - 23) | 10  (5 - 18) | 1  (0 - 2) |
|  | **Percent Outcomes Averted*** | 55%  (39 - 92%) | 54%  (38 - 91%) | 37%  (21 - 56%) | 27%  (12 - 49%) | 2%  (1 - 7%) |
| *Total  Population* | **AGE Cases** | 4980000  (2900000 - 9900000) | 4890000  (2670000 - 9480000) | 3260000  (1810000 - 6290000) | 2350000  (1230000 - 4870000) | 190000  (90000 - 660000) |
|  | **Outpatient Visits** | 695000  (339000 - 1470000) | 683000  (320000 - 1430000) | 458000  (213000 - 931000) | 328000  (146000 - 704000) | 14000  (-30000 - 71000) |
|  | **Hospitalizations** | 24200  (13500 - 63000) | 23700  (12500 - 59600) | 15800  (8300 - 36500) | 11300  (5600 - 28900) | 700  (200 - 3500) |
|  | **Deaths** | 213  (75 - 1011) | 208  (65 - 899) | 135  (47 - 520) | 97  (34 - 410) | 7  (2 - 54) |
|  | **Percent AGE Cases Averted** | 25%  (9 - 90%) | 25%  (9 - 89%) | 17%  (5 - 49%) | 12%  (3 - 44%) | 1%  (0 - 6%) |
|  | **Percent Outpatient Visitis Averted** | 31%  (12 - 91%) | 30%  (11 - 89%) | 20%  (7 - 50%) | 15%  (4 - 44%) | 1%  (-1 - 5%) |
|  | **Percent Hospitalizations Averted** | 18%  (6 - 89%) | 17%  (6 - 88%) | 12%  (3 - 45%) | 8%  (2 - 40%) | 1%  (0 - 5%) |
|  | **Percent Deaths Averted** | 9%  (2 - 88%) | 9%  (1 - 87%) | 6%  (1 - 42%) | 4%  (1 - 36%) | 0%  (0 - 4%) |
| *Notes:* | *As clinical outcomes are scaled from AGE cases, the percent outcomes averted are the same across outcomes within an age group. For the total population, individual percent outcomes averted are presented for each clinical outcome. | | | | | |

**Table 4: Vaccine efficiency by vaccination scenario (95% uncertainty interval based on LHS sensitivity analysis) by age group with no maternal antibody interference with vaccine efficacy and 90% vaccination coverage.** Vaccine efficiency measures are presented for the cumulative doses or vaccinated individuals and cumulative cases averted over five years.

| *Efficiency Measure* | *Efficiency Value by Vaccination Scenario  (95% UI)* | | | | |
| --- | --- | --- | --- | --- | --- |
| Doses to avert a case | 2.2  (1.3 - 4.2) | 2.2  (1.3 - 4.1) | 2.1  (1.2 - 3.9) | 2.8  (1.4 - 5.2) | 16.6  (5.2 - 35.4) |
| Vaccinees to avert a case | 0.8  (0.5 - 1.5) | 0.7  (0.4 - 1.4) | 1.1  (0.6 - 2.1) | 1.5  (0.8 - 2.9) | 16.6  (5.2 - 35.4) |
| Cases averted per dose | 0.5  (0.2 - 0.8) | 0.5  (0.2 - 0.8) | 0.5  (0.3 - 0.8) | 0.4  (0.2 - 0.7) | 0.1  (0 - 0.2) |
| Cases averted per vaccinee | 1.3  (0.7 - 2.2) | 1.3  (0.7 - 2.2) | 0.9  (0.5 - 1.6) | 0.7  (0.3 - 1.3) | 0.1  (0 - 0.2) |

**Table 5: Total clinical outcomes averted annually by vaccination scenario (95% uncertainty interval based on LHS sensitivity analysis) by age group with maternal antibody interference with vaccine efficacy and 90% vaccination coverage.**

| *Age Group* | *Clinical Outcome* | *Outcomes Averted by Vaccination Scenario  (95% SI)* | | | | |
| --- | --- | --- | --- | --- | --- | --- |
|  |  | **0/2/4m** | **2/4/6m** | **6/12m** | **12/24m** | **48m** |
| *Young Children (<5yrs)* | **AGE Cases** | 610000  (320000 - 1410000) | 1040000  (560000 - 2390000) | 1190000  (640000 - 2380000) | 1080000  (540000 - 2220000) | 100000  (50000 - 300000) |
|  | **Outpatient Visits** | 102000  (44000 - 263000) | 175000  (75000 - 453000) | 199000  (88000 - 465000) | 182000  (72000 - 423000) | 17000  (7000 - 55000) |
|  | **Hospitalizations** | 2600  (1400 - 6200) | 4500  (2400 - 10400) | 5100  (2700 - 10400) | 4600  (2300 - 9600) | 400  (200 - 1300) |
|  | **Deaths** | 4  (2 - 9) | 6  (3 - 15) | 7  (4 - 15) | 7  (3 - 14) | 1  (0 - 2) |
|  | **Percent Outcomes Averted*** | 10%  (6 - 19%) | 18%  (10 - 33%) | 20%  (9 - 38%) | 18%  (7 - 39%) | 2%  (1 - 6%) |
| *Total  Population* | **AGE Cases** | 820000  (440000 - 1960000) | 1410000  (760000 - 3490000) | 1670000  (880000 - 3860000) | 1560000  (760000 - 3600000) | 170000  (80000 - 630000) |
|  | **Outpatient Visits** | 114000  (49000 - 301000) | 200000  (88000 - 524000) | 234000  (101000 - 579000) | 216000  (83000 - 535000) | 14000  (-20000 - 69000) |
|  | **Hospitalizations** | 3800  (2000 - 10700) | 6600  (3300 - 19300) | 7900  (3800 - 22500) | 7500  (3300 - 22400) | 700  (100 - 3500) |
|  | **Deaths** | 26  (10 - 135) | 47  (17 - 243) | 61  (21 - 286) | 61  (19 - 308) | 6  (2 - 52) |
|  | **Percent AGE Cases Averted** | 4%  (1 - 15%) | 7%  (2 - 28%) | 8%  (2 - 32%) | 8%  (2 - 34%) | 1%  (0 - 6%) |
|  | **Percent Outpatient Visitis Averted** | 5%  (1 - 15%) | 9%  (3 - 28%) | 10%  (3 - 33%) | 10%  (2 - 35%) | 1%  (-1 - 5%) |
|  | **Percent Hospitalizations Averted** | 3%  (1 - 13%) | 5%  (1 - 24%) | 6%  (2 - 28%) | 5%  (1 - 31%) | 0%  (0 - 4%) |
|  | **Percent Deaths Averted** | 1%  (0 - 10%) | 2%  (0 - 20%) | 3%  (0 - 24%) | 3%  (0 - 28%) | 0%  (0 - 4%) |
| *Notes:* | *As clinical outcomes are scaled from AGE cases, the percent outcomes averted are the same across outcomes within an age group. For the total population, individual percent outcomes averted are presented for each clinical outcome. | | | | | |

**Table 6: Vaccine efficiency by vaccination scenario (95% uncertainty interval based on LHS sensitivity analysis) by age group with maternal antibody interference with vaccine efficacy and 90% vaccination coverage.** Vaccine efficiency measures are presented for the cumulative doses or vaccinated individuals and cumulative cases averted over five years.

| *Efficiency Measure* | *Efficiency Value by Vaccination Scenario  (95% UI)* | | | | |
| --- | --- | --- | --- | --- | --- |
|  | **0/2/4m** | **2/4/6m** | **6/12m** | **12/24m** | **48m** |
| Doses to avert a case | 13.3  (5.9 - 25.5) | 7.5  (3.4 - 14.1) | 4.0  (1.9 - 7.7) | 4.0  (1.8 - 7.9) | 18.7  (5.5 - 40.7) |
| Vaccinees to avert a case | 4.5  (2 - 8.8) | 2.5  (1.2 - 4.9) | 2.1  (1 - 4.1) | 2.1  (1 - 4.4) | 18.7  (5.5 - 40.7) |
| Cases averted per dose | 0.1  (0 - 0.2) | 0.1  (0.1 - 0.3) | 0.2  (0.1 - 0.5) | 0.2  (0.1 - 0.6) | 0.1  (0 - 0.2) |
| Cases averted per vaccinee | 0.2  (0.1 - 0.5) | 0.4  (0.2 - 0.9) | 0.5  (0.2 - 1) | 0.5  (0.2 - 1) | 0.1  (0 - 0.2) |
